# Lipidome-wide analysis of concurrent and longitudinal blood pressure patterns in school-aged children

**DOI:** 10.64898/2026.11.09.26367552

**Authors:** Rossella Alfano, Tim S Nawrot, Kathrin Barth, Nick Giesberts, Eleni Renaers, Maartje Vangeneugden, Michelle Plusquin

## Abstract

**Background:** Elevated blood pressure (BP) in childhood is associated with cardiovascular risk, but its molecular lipid correlates remain poorly characterized. We investigated the association of plasma lipid species and patterns with concurrent and longitudinal BP phenotypes in school-aged children.

**Methods:** In the Belgian ENVIRONAGE prospective birth-cohort, 195 children aged 9 to 12 years were examined between July 2021 and September 2024. A total of 1,232 plasma lipid species were quantified using liquid chromatography-electrospray ionization-tandem mass spectrometry. Associations with systolic and diastolic BP percentiles were examined using multivariable linear regression with permutation-based family-wise error rate (FWER) correction. Lipid-set enrichment analysis, with P values from sample permutation to account for correlation among lipid species, evaluated coordinated associations. Significant findings were examined across longitudinal BP patterns in 177 children with BP measured at ages 4-6 and 9-12.

**Findings:** Each 1-SD increase in triglyceride (TG) 58:9 fatty acyl (FA) 22:4 was associated with 7.36 higher systolic BP percentile points (95% CI=3.91-10.80; *P*_FWER_=0.03). TG 58:9 FA 22:4 was associated with incident (OR=2.78; 95% CI=1.33-5.79) and persistent (OR=2.14; 95% CI=1.22-3.77), but not resolved elevated BP vs stable-normal BP. Among lipid sets, species with a total carbon number of 38, mainly PC and PE, were inversely enriched with diastolic BP percentile.

**Interpretation:** Childhood BP was associated with higher levels of a highly polyunsaturated long-chain TG, closely related to current BP elevation and not elevated among children whose BP had normalized.

**Funding:** Special Research Fund, Flemish Scientific Research Fund, Methusalem and Kom op Tegen Kanker.

**Research in context:** *Evidence before this study:* We searched PubMed for studies published from inception to 13 October 2026, without language restrictions, for human studies, using the following combinations of terms: (“child*” OR “adolescen*”) AND (“blood pressure” OR “hypertension”) AND (“lipidomic*” OR “lipidome”). Previous pediatric studies have implicated glycerophospholipids, sphingolipids and triglycerides, but most were conducted in specific clinical, dietary, environmental, or ethnic settings. We found little population-based evidence from comprehensive lipidome-wide analyses and no studies examining whether lipidomic signatures differ according to longitudinal blood pressure patterns spanning early and later childhood.

*Added value of this study:* In a population-based birth cohort, we measured 1,232 lipid species in plasma from 195 children aged 9-12 years and related them to blood pressure at that age and at ages 4-6 years. One long-chain, highly unsaturated triglyceride was associated with higher systolic blood pressure. The triglyceride association was present in children whose blood pressure became or remained elevated, but not in those whose elevated blood pressure had normalized. Accounting for the correlation among lipids, only one lipid group, phospholipids with 38 carbon atoms, was inversely enriched with diastolic blood pressure.

*Implications of all the available evidence:* Together with adult studies linking triglyceride species to hypertension, our findings suggest that a triglyceride species is also associated with blood pressure in children. The lead triglyceride species was more closely related to current elevation, and was not detectable in children with normal current blood pressure but earlier elevation. Smaller signatures of earlier blood pressure cannot be excluded, studies with repeated lipidomics and blood pressure assessment from early childhood are needed to determine whether earlier blood pressure leaves a more subtle trace in the lipidome.

## Introduction

Elevated blood pressure (BP) is a major modifiable contributor to cardiovascular disease worldwide [1] and sustained hypertension already affects about 4% of children and adolescents globally [2]. Childhood BP is associated with subclinical cardiovascular alterations [3], tracks into adulthood [4], and is prospectively associated with cardiovascular events later in life [5, 6]. Its relevance may depend on its course: within childhood, incident and persistent elevated BP were associated with greater left ventricular mass, whereas elevated BP that resolved was not [7]. Identifying molecular signatures of BP and its longitudinal patterns during childhood could help characterize early biological pathways underlying BP regulation [8].

Elevated BP frequently co-occurs with dyslipidemia [9]. Lipidomics, which quantifies hundreds to thousands of individual lipid species, can resolve signals that standard lipid measures cannot. In adults, diacylglycerols (DGs), triacylglycerols (TGs), phosphatidylcholines (PCs), and phosphatidylethanolamines (PEs) have been associated with BP and hypertension, cross-sectionally and longitudinally, although the species and direction of association vary between populations [10–16]. Pediatric lipidomic-BP studies have implicated glycerophospholipids, sphingolipids and TGs but have largely focused on selected populations, including children with overweight or obesity [17], or on specific dietary [18, 19], environmental [20], or ethnic contexts [21]. Although pediatric longitudinal metabolomic-BP studies exist [22], none has examined whether lipidomic profiles differ by the course of BP over childhood.

We investigated associations of 1,232 plasma lipid species with systolic and diastolic BP in 195 children aged 9-12 years participating in the ENVIRONAGE birth cohort. We additionally tested whether signals clustered by lipid class, total carbon number, and degree of unsaturation. In 177 children with BP measured at both the 4-6-year and 9-12-year childhood follow-up visits, we explored whether BP-associated lipid species and classes differed across stable-normal, incident, persistent, and resolved elevated longitudinal BP patterns.

## Methods

The study was reported in accordance with Strengthening the Reporting of Observational Studies in Epidemiology (STROBE) guidelines.

### Study population

The ongoing ENVIRonmental influence ON AGEing in early life (ENVIRONAGE) study is a population-based prospective birth cohort established in 2010 in Limburg, Belgium [23]. Mother-newborn pairs are recruited at birth at East-Limburg Hospital in Genk, Belgium, and are subsequently invited to participate in two follow-up examinations later in life (between 4 and 6 years of age and between 9 and 12 years of age). At enrolment, eligibility required mothers to be able to complete questionnaires in Dutch and to have not planned a caesarean section. The ENVIRONAGE study was conducted in accordance with the principles of the Declaration of Helsinki and was approved by the ethics committees of Hasselt University and East-Limburg Hospital (Genk, Belgium; reference nos. B371201216090, B371201524537, and B1152021000007). Written informed consent was obtained from the mothers at cohort enrolment. For the childhood follow-ups, written informed consent was obtained from the parents or legal guardians, and assent was obtained from the participating children.

The present study includes 195 children participating between July 2021 and September 2024 in the second childhood follow-up examination at 9-12 years of age, who provided a blood sample for lipidomic profiling and had a valid BP measurement. 177 of these children also had a valid BP measurement at the first follow-up examination at 4-6 years of age. The number of participants included in individual statistical models depended on covariate availability, as specified in the Results.

### Lipidomics

During the follow-up examination at 9-12 years of age, venous blood was collected by a trained fieldworker. Plasma was separated and stored at −80°C until lipidomic analysis.

Lipidomic analyses were performed at the Lipometrix lab at KU Leuven (Leuven, Belgium) using liquid chromatography electrospray ionization tandem mass spectrometry (LC-ESI-MS/MS) on a Nexera X2 UHPLC system (Shimadzu) coupled with hybrid triple quadrupole/linear ion trap mass spectrometer (6500+ QTRAP system; AB SCIEX). Lipids were quantified by scheduled multiple-reaction monitoring using internal standard signals and adhered to the guidelines of the Lipidomics Standards Initiative (LSI) (level 2 type quantification as defined by the LSI). Detailed extraction procedures, chromatographic conditions, mass-spectrometry settings, and lipid-class-specific transitions are provided in the **Supplemental Methods**.

The analytical platform initially measured 2,017 lipid species. Lipid nomenclature was standardized using the *rgoslin* R package. Preprocessing was performed in R, version 4.5.0. Lipid species with missing measurements in more than 35% of participants were excluded. This filtering removed 785 lipids, leaving 1,232 lipids for analysis. Among the retained lipids, 8,605 measurements (approximately 3.6% of the lipidomic data matrix) were missing and were imputed using the correlation-based k-nearest-neighbour method (kNN-CR), selected on the basis of previous simulation evidence for lipidomic data [24]. Lipid values were subsequently log₂-transformed to reduce right skewness. For lipid-wide association analyses, the log₂-transformed values were standardized to a mean of zero and a standard deviation of one.

### Blood pressure assessment

During the follow-up examination at 9-12 years of age, child BP was measured with an automated upper-arm blood-pressure monitor (Omron 705IT; Omron Corporation), equipped with a cuff adapted to the arm size of children. Measurements were performed according to the European Society of Hypertension guideline [25]. After 5 minutes of rest in supine position, a trained observer obtained 5 consecutive readings of the systolic BP and diastolic BP with 1-minute intervals. Mean systolic BP and diastolic BP were based on the mean of the last 3 readings. Mean arterial pressure (MAP) was calculated as diastolic BP plus 40% of pulse pressure (the difference between systolic BP and diastolic BP) [26]. Elevated BP and hypertension stages were defined according to the 2017 American Academy of Pediatrics (AAP) guideline [27]. Age-, sex-, and height-specific BP percentiles were calculated for systolic BP and diastolic BP via the online calculator (https://bsolomon.us/app/pediatric-bp/).

For a subset of children (N=177), BP during early childhood was measured during the follow-up visit at 4-6 years of age using the same methodology.

### Longitudinal BP patterns

Based on BP status at the first (4-6 years) and second (9-12 years) follow-up, children were classified into four categories: stable normal, defined as normal BP at both visits; incident elevated BP, defined as normal BP at 4-6 years and elevated BP or hypertension at 9-12 years; persistent elevated BP, defined as elevated BP or hypertension at both visits; and resolved elevated BP, defined as elevated BP or hypertension at 4-6 years and normal BP at 9-12 years.

### Covariates

For the primary cross-sectional lipidomic analysis of BP at 9-12 years, potential confounders were selected a priori based on hypothesized causal relationships represented in a directed acyclic graph (DAG; **Figure S1**) and included child age, sex, ethnicity, height, weight, highest parental educational attainment, household smoking, physical activity, screen time, and season of examination. Time of blood collection and plasma storage duration were additionally included as temporal and preanalytical covariates, respectively. Pubertal development, ambient temperature, fish and seafood consumption, cardiometabolic conditions, corticosteroid use, and overweight/obesity were evaluated in sensitivity analyses. Detailed definitions and methods of collection at the visit at 9-12 years are provided in the **Supplemental Methods**.

### Statistical analysis

All data management and statistical analyses were done using R version 4.5.0. Participant characteristics were presented as means (standard deviation, SD) or frequency (percentage, %). All statistical tests were two-sided. Analyses were conducted on complete cases for the covariates of each model.

Because sample size was determined by cohort participation, we performed a simulation-based analysis to estimate the minimum detectable effect (MDE) as detailed in the **Supplemental Methods.**

The analysis workflow is depicted in **Figure 1**. Primary analyses comprised cross-sectional lipidome-wide association analyses of BP at 9-12 years, followed by LSEA; secondary analyses evaluated whether significant species and enriched lipid classes were also associated with longitudinal BP patterns between 4-6 and 9-12 years.

**Figure 1:**
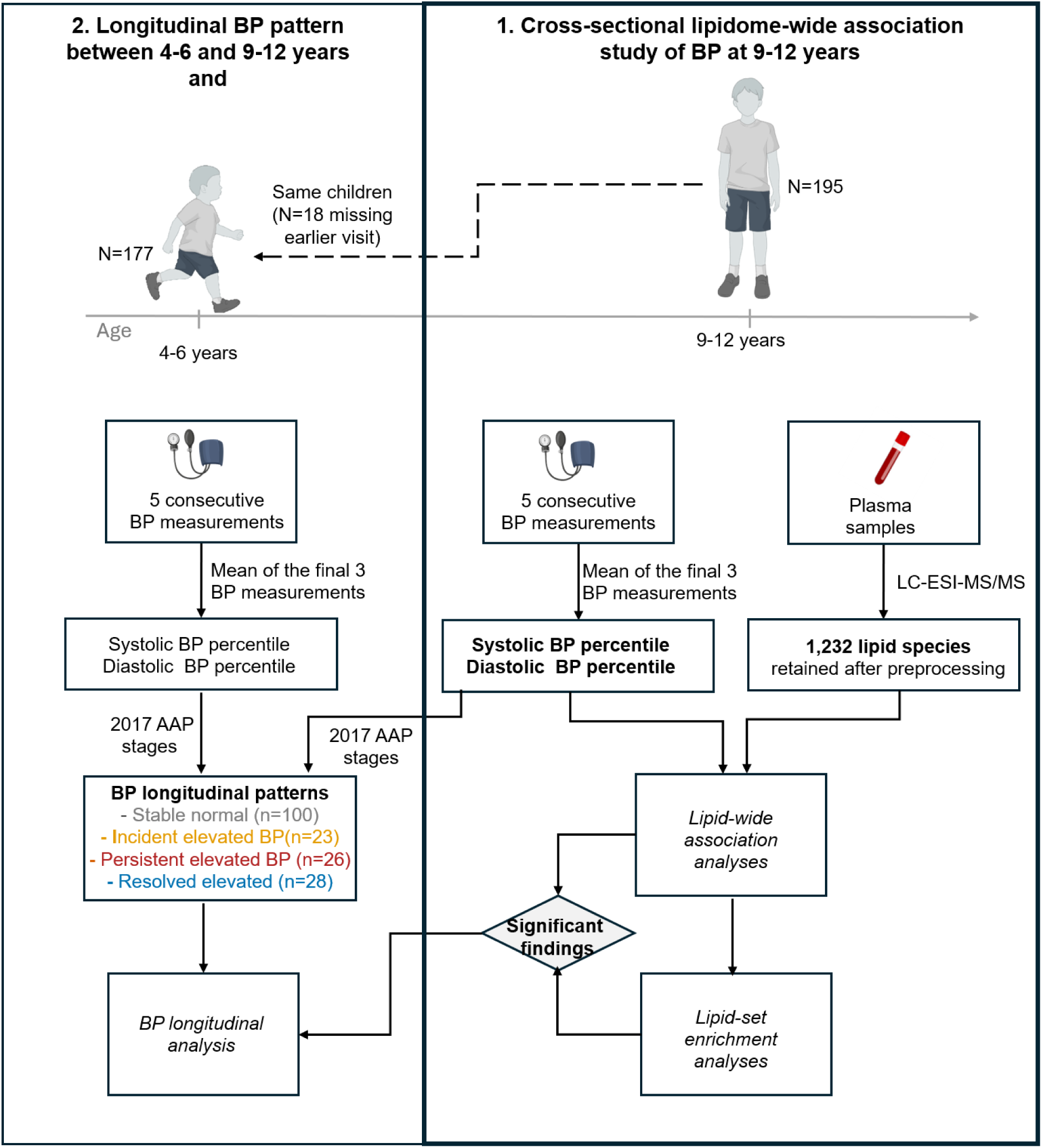
Study design and analytical workflow. (1) Blood pressure (BP) was measured and plasma lipid species were quantified in 195 children aged 9-12 years participating in the second childhood follow-up of the ENVIRonmental influence ON AGEing in early life (ENVIRONAGE) birth cohort. BP was measured five times using an automated upper-arm BP monitor; the mean of the final three measurements was used to calculate systolic and diastolic BP percentiles. Plasma lipid species were quantified by liquid chromatography-electrospray ionization-tandem mass spectrometry (LC-ESI-MS/MS); of 2,017 quantified lipid species, 1,232 passed predefined quality-control criteria and were retained for cross-sectional lipidome-wide association analyses and lipid-set enrichment analysis (LSEA). Lipid species meeting the lipidome-wide significance threshold and lipid classes showing significant enrichment were identified as “significant findings”. (2) 177 of the 195 children (91%) also had valid BP measured at the first childhood follow-up visit (4-6 years). At both visits, normal BP, and elevated BP or hypertension were defined according to the 2017 American Academy of Pediatrics (AAP) guidelines. Four longitudinal BP patterns were identified based on AAP classification at both visits: stable normal BP (normal BP at both visits, n=100), incident elevated BP (normal BP at 4-6 years, elevated BP or hypertension at 9-12 years, n=23), persistent elevated BP (elevated BP or hypertension at both visits, n=26), and resolved elevated BP (elevated BP or hypertension at 4-6 years, normal BP at 9-12 years, n=28). Lipid species meeting the lipidome-wide significance threshold in the cross-sectional analysis and lipid classes showing significant enrichment in LSEA (described in 1 as “significant findings”) were evaluated in relation to longitudinal BP pattern. AAP=American Academy of Pediatrics; BP=blood pressure; LC-ESI-MS/MS=liquid chromatography-electrospray ionization-tandem mass spectrometry; LSEA=lipid-set enrichment analysis.

### Cross-sectional lipidome-wide association analysis

Cross-sectional lipidome-wide associations with BP at 9-12 years were examined using multivariable linear regression. Separate models were fitted for each individual lipid species and BP outcome. Systolic and diastolic BP percentiles were considered the primary outcomes. Systolic BP, diastolic BP, and MAP, expressed in mmHg, were considered secondary outcomes. Models were adjusted for the confounders identified in the DAG (**Figure S1**): child sex, age, height, weight, ethnicity, season, parental education, household smoking, physical activity and screen time, along with time of blood collection and plasma storage duration, which were added as temporal and preanalytical covariates. Regression coefficients and their 95% confidence intervals represent the difference in BP percentile points, or in mmHg for the secondary outcomes, associated with a 1-standard-deviation higher log₂-transformed lipid level. To account for multiple testing while preserving correlation among lipid species, empirical permutation-based family-wise error rate (FWER) p-values (*P*_FWER_) were obtained using a single-step maximum-statistic permutation procedure with 50,000 Freedman-Lane residual permutations, as detailed in **Supplemental Methods** [28–31]. Lipidome-wide significance was defined as *P*_FWER_<0.05. Quantile-quantile plots were used to compare the observed and expected distributions of nominal P values for each lipid-wide analysis. Test-statistic inflation factors (λ) were calculated as the ratio of the median observed 1-degree-of-freedom chi-square statistic to its expected median under the null distribution. The null distribution of the genomic inflation factor λ was obtained from the same Freedman-Lane permutations.

For lipid associations with *P*_FWER_<0.05, potential departure from linearity and model assumptions were checked as described in the **Supplemental Methods**. As exploratory secondary analyses, distributions of these lipid species were visualized across pediatric BP categories and associations with BP category were examined using multinomial logistic regression adjusted for the same covariates of the primary model. To assess linear trend, BP category was additionally modelled as an ordinal outcome and the P value for the lipid term was used as the test for trend. Nominal P<0.05 was used for these targeted analyses.

### Lipid-set enrichment analysis

LSEA was performed using the *lipidr* package to determine whether coordinated associations with BP occurred within lipid sets defined by lipid class, total carbon number, or degree of unsaturation. For each primary and secondary BP outcome, lipid species were ranked according to the signed t-statistic from the corresponding model. Positive and negative normalized enrichment scores (NES) indicated lipid sets tending towards positive and inverse associations with BP, respectively. Lipid sets containing fewer than five measured species were not evaluated. Enrichment was assessed using the fast gene-set enrichment algorithm implemented through fgsea. Because *lipidr*’s preranked procedure permutes lipid labels, which assumes independence between species, P values were obtained by permuting BP residuals across children (Freedman-Lane, 50,000 permutations). Benjamini-Hochberg false discovery rate (FDR)-adjusted *P* was calculated separately for each BP outcome and set type (lipid class, total carbon number, degree of unsaturation), and FDR-adjusted *P*<0.05 for was considered evidence of lipid-set enrichment.

### Sex interaction

Potential effect modification by sex was examined in exploratory analyses by adding a product term between each standardized lipid concentration and sex to the primary model. To account for pubertal development, we also tested the interaction model additionally adjusting for the Pubertal Development Scale (PDS) score.

### Longitudinal BP patterns analysis

In the subset of 177 children with BP data available at both childhood follow-up visits, we evaluated whether lipid species meeting the lipidome-wide significance threshold and lipid classes showing significant enrichment in the cross-sectional analyses were also associated with longitudinal BP patterns. Associations of the selected lipid species with BP pattern were assessed using multinomial logistic regression, with stable normal BP as the reference category. To compare the longitudinal groups directly, the multinomial models were repeated with resolved elevated BP as the reference category (incident vs resolved; persistent vs resolved), and persistent elevated BP as the reference for incident vs persistent. Models were adjusted using the same covariate sets as the cross-sectional models and nominal P<0.05 was considered significant. LSEA was repeated using the longitudinal BP-pattern association statistics ranking lipid species by the Wald t-statistic, with interpretation restricted to lipid classes that showed significant enrichment in the primary cross-sectional analysis and FDR P <0.05 was considered significant.

### Sensitivity analyses

Sensitivity analyses of lipid species meeting the lipidome-wide significance threshold and lipid classes showing significant enrichment in the cross-sectional analyses were conducted by repeating the analyses by: (i) additionally adjusting for the PDS score; (ii) removing child weight from the adjustment; (iii) replacing, in the adjustment set, season of examination with average daily temperature on the day of examination among children with available meteorological information; (iv) additionally adjusting for fish and seafood consumption; (v) excluding children with reported cardiometabolic conditions (n=2); (vi) excluding children with reported corticosteroid use (n=5); and (vii) excluding children with overweight or obesity (n=35). In addition, the complete lipidome-wide and lipid-set enrichment analyses, including the longitudinal analyses, were repeated using a basic model including a reduced covariate set (child sex, age, height, ethnicity, season, time of blood collection and plasma storage duration), which does not require complete data on weight, parental education, household smoking, physical activity and screen time and therefore retains a larger sample (N = 195 cross-sectionally; N = 177 longitudinally). Robustness was evaluated based on the direction, magnitude, and precision of effect estimates relative to the primary analysis.

## Results

### Study population

Characteristics of the 195 ENVIR*ON*AGE participants in the study are presented in **Table 1**. At the 9-12-year follow-up, the children, of whom 54.9% were girls and most were of European descent (92.8%), had a mean (SD) age of 10.04 (0.82) years. Mean (SD) systolic BP, diastolic BP, and MAP were 107.00 (9.63), 64.70 (6.53), and 81.62 (6.83) mmHg, respectively. Mean (SD) systolic and diastolic BP percentiles were 68.68 (24.08) and 61.68 (20.58), respectively, with their distribution shown in **Figure 2A**. Based on the 2017 AAP classification, 142 children (72.8%) had BP within the normal range, whereas 20 (10.3%) had elevated BP, 23 (11.8%) had BP in the stage 1 hypertension range, and 10 (5.1%) in the stage 2 hypertension range. At the first childhood follow-up, conducted at 4-6 years of age, mean (SD) systolic and diastolic BP percentiles were 70.57 (22.90) and 60.02 (23.89) percentile points, respectively (**Figure S2**). BP classification at both childhood examinations was available for 177 children. Of these 177, 100 (56.5%) had normal BP at both time points (stable normal BP pattern), whereas 23 (13.0%) transitioned from normal BP at 4-6 years to elevated or hypertensive BP at 9-12 years (incident elevated BP pattern, **Figure 2B**). Elevated or hypertensive BP persisted across both examinations in 26 children (persistent elevated BP pattern, 14.7%), while 28 (15.8%) transitioned from elevated or hypertensive BP at 4-6 years to normal BP at 9-12 years (resolved elevated BP pattern). Characteristics of the study population by BP patterns are reported in **Table S1**.

**Figure 2:**
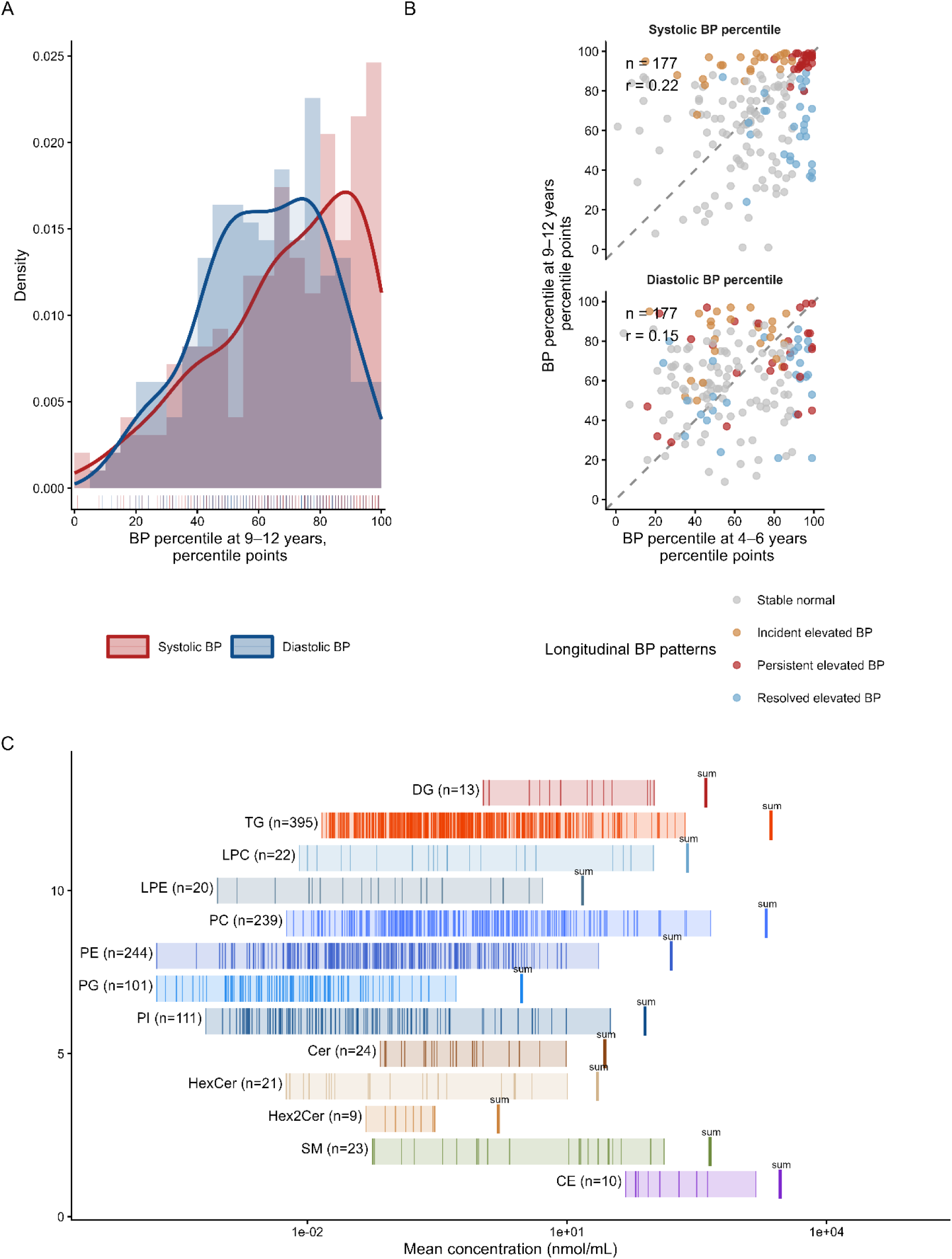
Blood pressure distribution at 9-12 years, longitudinal BP tracking between 4-6 and 9-12 years and plasma lipidomic coverage at 9-12 years. **(A)** Distributions of systolic (red) and diastolic (blue) blood pressure (BP) percentiles at 9-12 years (N=195). Bars and solid lines represent histograms and kernel density estimates, respectively; rug marks represent individual observations. **(B).** Patterns of systolic and diastolic BP percentiles between the first (4-6 years, x-axis) and second (9-12 years, y-axis) childhood follow-up visits, among the 177 children with BP measured at both visits. Each point represents one child, coloured according to longitudinal BP pattern: stable normal (grey), incident elevated BP (orange), persistent elevated BP (red), and resolved elevated BP (blue). Normal BP, elevated BP, and hypertension were classified according to the 2017 American Academy of Pediatrics (AAP) guideline. Pearson correlation coefficients (r) between BP percentiles at the two visits are reported. **(C)** Coverage and concentration range of the retained plasma lipidome at 9-12 years according to lipid class. Vertical lines represent the mean concentration (nmol/mL) of individual lipid species across participants, calculated from the original non-imputed measurements. Shaded areas represent the range of mean species concentrations within each lipid class. The number of retained species is shown next to each class. The “sum” mark indicates the summed mean concentration of all retained species within a class. Concentrations are displayed on a logarithmic scale. AAP=American Academy of Pediatrics; BP=blood pressure; CE= cholesteryl ester; Cer= ceramide; DG= diacylglycerol; HexCer= hexosylceramide; Hex2Cer= dihexosylceramide; LPC= lysophosphatidylcholine; LPE= lysophosphatidylethanolamine; PC= phosphatidylcholine; PE= phosphatidylethanolamine; PG= phosphatidylglycerol; PI= phosphatidylinositol; SM= sphingomyelin; TG= triacylglycerol.

**Table 1:** Characteristics of 195 ENVIR*ON*AGE participants.

| **Characteristics** | **Overall (N=195)** | **Missing, n (%)** |
|---|---|---|
| **9-12 years** |  |  |
| Age, years | 10.04 (0.82) | 0 (0.0) |
| Child sex, girls | 107 (54.9) | 0 (0.0) |
| Ethnicity, non-European | 14 (7.18) | 0 (0.0) |
| Weight, kg | 35.59 (7.73) | 1 (0.5) |
| Height, cm | 141.20 (7.49) | 0 (0.0) |
| BMI, kg/m² | 17.70 (2.68) | 1 (0.5) |
| Overweight or obese | 35 (17.9) | 1 (0.5) |
| Highest parental educational attainment, low | 38 (19.5) | 5 (2.6) |
| Household smoking, yes | 42 (21.5) | 8 (4.1) |
| Average daily screen time, h/day | 3.59 (2.17) | 8 (4.1) |
| Average daily physical activity, h/day | 4.09 (2.36) | 8 (4.1) |
| Pubertal Development Scale score | 1.34 (0.45) | 10 (5.1) |
| Season of examination |  | 0 (0.0) |
| Winter | 46 (23.6) |  |
| Spring | 45 (23.1) |  |
| Summer | 63 (32.3) |  |
| Autumn | 41 (21.0) |  |
| Average daily temperature, °C | 11.94 (5.93) | 35 (17.9) |
| Fish and seafood consumption, time per week | 1.87 (1.63) | 31 (15.9) |
| Time of blood collection, hour of day | 12.31 (2.19) | 0 (0.0) |
| Plasma storage duration, years | 3.06 (0.93) | 0 (0.0) |
| Systolic BP, mmHg | 107.00 (9.63) | 0 (0.0) |
| Diastolic BP, mmHg | 64.70 (6.53) | 0 (0.0) |
| Mean arterial pressure, mmHg | 81.62 (6.83) | 0 (0.0) |
| Systolic BP percentile, percentile points | 68.68 (24.08) | 0 (0.0) |
| Diastolic BP percentile, percentile points | 61.68 (20.58) | 0 (0.0) |
| BP category |  | 0 (0.0) |
| Normal BP | 142 (72.8) |  |
| Elevated BP | 20 (10.3) |  |
| Stage 1 hypertension | 23 (11.8) |  |
| Stage 2 hypertension | 10 (5.1) |  |
| **4-6 years** |  |  |
| Systolic BP percentile, percentile points | 70.57 (22.90) | 18 (9.2) |
| Diastolic BP percentile, percentile points | 60.02 (23.89) | 18 (9.2) |
| **Longitudinal BP patterns (BP category transitions between 4-6 years and 9-12 years)** |  |  |
| Stable-normal BP | 100 (51.3) | 18 (9.2) |
| Resolved elevated BP | 28 (14.4) |  |
| Incident elevated BP | 23 (11.8) |  |
| Persistent elevated BP | 26 (13.3) |  |

In plasma samples from the 9-12-year-old children, following preprocessing, 1,232 lipid species measured across 13 lipid classes were retained for further analysis. TGs constituted the largest class (n=395), followed by PEs (n=244) and PCs (n=239) (**Figure 2C**). Mean species concentrations spanned approximately six orders of magnitude, from 1.81 × 10^−04^ to 1522 nmol/mL.

Complete data on all covariates of the primary model were available for 186 children (and 173 of the 177 children with longitudinal BP data). In this population, the lipidome-wide association study was estimated to have MDE of β≈8.68 systolic BP percentile points and β≈7.46 diastolic BP percentile points per 1-SD higher lipid concentration (**Figure S3**).

### Cross-sectional BP and lipidomics at 9-12 years

#### Lipidome-wide analyses

For systolic BP percentile, TG 58:9 FA 22:4 was the only lipid species meeting the lipidome-wide FWER threshold. In the primary model (N=186), upon adjustment for child weight, average daily physical activity, average daily screen time, highest parental educational attainment, and household smoking, a 1-SD higher log₂-transformed TG 58:9 FA 22:4 concentration was associated with 7.36-point higher systolic BP percentile (95% CI= 3.91 to 10.80; *P* = 3.98 × 10^−05^; *P*_FWER_=0.03) (**Figure 3A**, **Table S2**). No lipid species passed the FWER threshold for diastolic BP percentile (**Figure 3B**), or for systolic, diastolic BP or MAP expressed in mmHg (**Figure S4D-F**, **Table S2**). Quantile-quantile plots showed the largest departure from the expected P-value distribution for systolic BP percentile (λ=2.45), whereas the distributions for the remaining BP outcomes were closer to expectation (e.g., λ=1.07 for diastolic BP percentile) (**Figure S5**). The observed value of λ for systolic BP percentile lay at the upper end of its null distribution derived from the Freedman-Lane permutations (median=0.96; 95th percentile=2.31; permutation P=0.04), indicating that the apparent inflation largely reflected inter-lipid correlation, with borderline evidence of a diffuse positive shift (mean t=0.89). Diagnostic analyses did not materially alter the association of TG 58:9 FA 22:4 with systolic BP percentile (**Figure S6**) and restricted cubic spline analysis provide limited evidence of departure from linearity (P for nonlinearity=0.05, **Figure 3C**). Each 1-SD higher TG 58:9 FA 22:4 was associated with higher odds of stage 1 hypertension vs normal BP (OR per 1-SD lipid= 3.03; 95% CI= 1.58 to 5.82; P=8.51×10^−04^); test for linear trend across ordered BP categories supported a gradient across BP categories (OR per 1-SD lipid=2.25; 95% CI= 1.49 to 3.39; P for trend=3.48×10^−05^) (**Figure 3D**).

**Figure 3:**
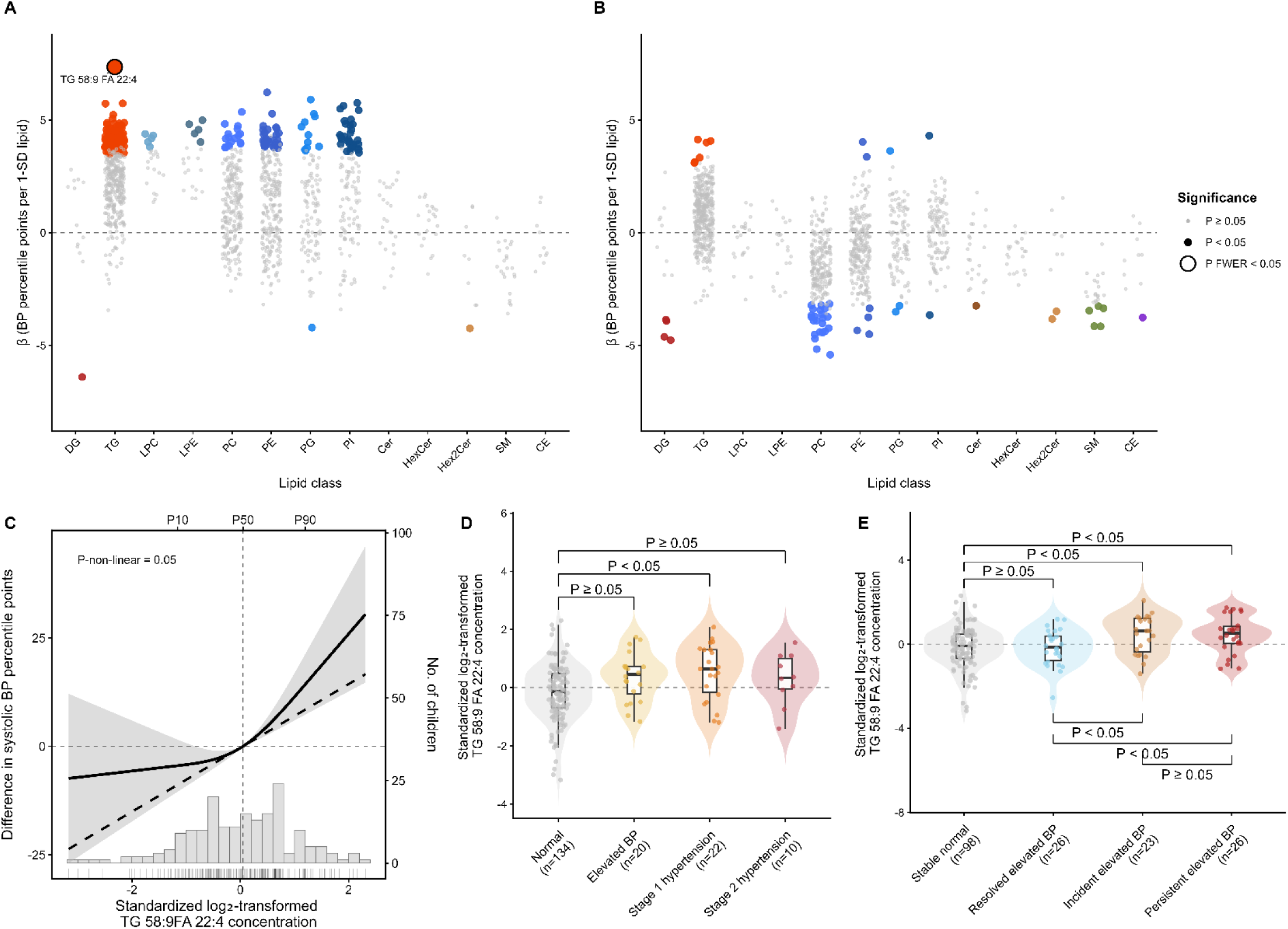
Lipidome-wide associations with blood pressure at 9-12 years and further characterization of the lead finding, TG 58:9 FA 22:4, including its longitudinal association with blood pressure patterns between 4-6 to 9-12 years. Regression coefficients from primary models of cross-sectional lipidome-wide association studies of **(A)** systolic and **(B)** diastolic blood pressure (BP) percentiles at 9-12 years (N=186), presented according to lipid class. Coefficients represent the difference in BP percentile points associated with a 1-standard-deviation higher log₂-transformed lipid concentration. Grey points represent nominal P≥0.05; small coloured points represent nominal P<0.05; enlarged coloured points with a black outline represent significance based on family-wise error rate-adjusted p-value (P_FWER_) <0.05. Lipids with P_FWER_ <0.05 are labelled. The horizontal dashed line indicates the null value. **(C)** Restricted cubic spline association between TG 58:9 FA 22:4 and systolic BP percentile at 9-12 years in the primary model (N=186). The solid line and shaded area represent the estimated difference in systolic BP percentile points and its 95% confidence interval from a restricted cubic spline model with three knots; the dashed line represents the corresponding linear association. Estimates are expressed relative to the median TG 58:9 FA 22:4 concentration, indicated by the vertical dashed line. The histogram and rug marks show the distribution of standardized log₂-transformed TG 58:9 FA 22:4 concentrations among the 186 children included in the analysis. P10, P50, and P90 indicate the 10th, 50th, and 90th percentiles. P-non-linear presents the Wald test of the nonlinear spline component. The violin plots visualize standardized log₂-transformed TG 58:9 FA 22:4 concentrations according to **(D)** pediatric BP category (normal BP, elevated BP, stage 1 hypertension, stage 2 hypertension) defined according to the 2017 American Academy of Pediatrics guideline in the study population at 9-12 years (N=186), and **(E)** longitudinal BP patterns (stable normal, incident elevated BP, persistent elevated BP, resolved elevated BP) in the subset of 173 children with BP measured at both 4-6 and 9-12 follow-up visits. Brackets indicate targeted multinomial logistic regression contrasts; labels indicate nominal P<0.05 or P≥0.05. All panels show results from models adjusted for child age, sex, height, weight, average daily physical activity, average daily screen time, ethnicity, highest parental educational attainment, household smoking, season of examination, time of blood collection, and plasma storage duration. CE= cholesteryl ester; Cer= ceramide; DG= diacylglycerol; HexCer= hexosylceramide; Hex2Cer= dihexosylceramide; LPC= lysophosphatidylcholine; LPE= lysophosphatidylethanolamine; PC= phosphatidylcholine; PE= phosphatidylethanolamine; PG= phosphatidylglycerol; PI= phosphatidylinositol; SM= sphingomyelin; TG= triacylglycerol.

#### LSEA

In the LSEA, one lipid set was associated with BP: species with a total carbon number of 38 were inversely associated with diastolic BP percentile (NES=−1.81; FDR=0.04), with a leading edge dominated by PC and PE carrying 20- and 22-carbon fatty acyl chains (**Figure 4**). No lipid class or unsaturation set reached significance; PC showed a borderline inverse enrichment with diastolic BP (FDR=0.06). Within the TG class specifically, TG 58:9 FA 22:4 was located within a broader region of predominantly positive associations among long-chain, polyunsaturated TGs (**Figure 4D**). Complete lipid-set enrichment results and leading-edge species are provided in **Table S3.**

**Figure 4:**
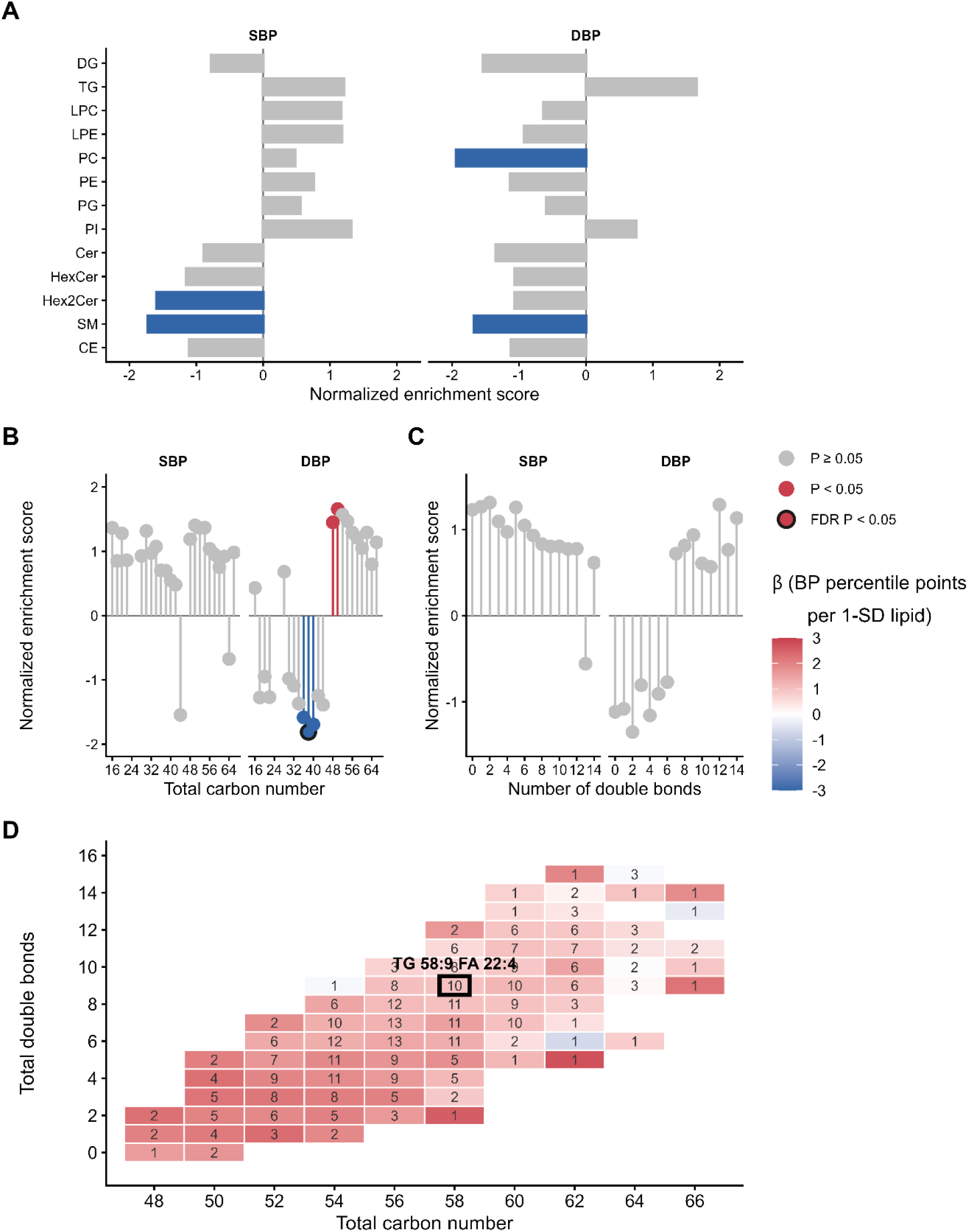
Lipid-class and structural enrichment of blood pressure at 9-12 years and structural distribution of triacylglycerol associations. (**A–C**) Lipid-set enrichment analysis (LSEA) of systolic (SBP) and diastolic blood pressure (DBP) percentiles at 9–12 years (N=186), for lipid sets defined by (A) lipid class, (B) total carbon number, and (C) total number of double bonds. Lipid species were ranked according to the signed *t*-statistic from the corresponding regression model. Bars and points represent normalized enrichment scores (NES), with positive and negative values indicating enrichment toward positive and inverse associations with BP, respectively. Grey indicates nominal *P*≥0.05; red and blue indicate nominal *P*<0.05 for positive and negative NES, respectively; a black outline indicates FDR-adjusted *P*<0.05. (**D**) Mean regression coefficients (β) for the association of triacylglycerol (TG) species with SBP percentile, grouped according to total carbon number and total number of double bonds. Cell colour represents the mean β, expressed as BP percentile points per 1-SD higher log₂-transformed lipid concentration, and numbers within cells indicate the number of TG species sharing that structural composition. The cell containing TG 58:9 FA 22:4, the only lipid species reaching lipidome-wide significance in the primary model, is outlined and labelled. All analyses were adjusted for child age, sex, height, weight, ethnicity, highest parental educational attainment, household smoking, average daily physical activity, average daily screen time, season of examination, time of blood collection, and plasma storage duration. CE= cholesteryl ester; Cer= ceramide; DBP=diastolic blood pressure; DG= diacylglycerol; HexCer= hexosylceramide; Hex2Cer= dihexosylceramide; LPC= lysophosphatidylcholine; LPE= lysophosphatidylethanolamine; PC= phosphatidylcholine; PE= phosphatidylethanolamine; PG= phosphatidylglycerol; PI= phosphatidylinositol; SBP=systolic blood pressure; SM= sphingomyelin; TG= triacylglycerol.

#### Sex interaction

We identified evidence of effect modification by sex for the association between PE 20:1 22:0 and diastolic BP percentile (βinteraction=−11.93 BP percentile points; 95% CI, −17.68 to −6.18; nominal P=6.52×10⁻⁵; P_FWER_=0.044) (**Figure S7**). The interaction remained similar after additional adjustment for pubertal development score (βinteraction=−11.92; 95% CI, −17.71 to −6.14; nominal P=7.31×10⁻⁵; P_FWER_=0.049) (**Table S4**). LSEA did not identified coordinated effect modification by sex across the lipid classes (**Figure S8**).

### Longitudinal BP patterns between 4-6 and 9-12 years and candidate lipid species and classes

Among 173 children with BP at 4-6 and 9-12 years and complete covariates, a 1-SD higher TG 58:9 FA 22:4 was associated with higher odds of incident (OR 2.78, 95% CI 1.33-5.79; P=6.47×10⁻^3^) and persistent elevated BP (OR 2.14, 1.22-3.77; P=8.02×10⁻^3^) compared with stable-normal BP, but not of resolved elevated BP (OR 0.94, 0.56-1.59; P=0.82). The lipid was higher in incident (OR 2.94, 1.27-6.85; P=0.01) and persistent (OR 2.27, 1.14-4.54; P=0.02) than in resolved elevated BP, and did not differ between incident and persistent elevated BP (OR 1.30, 0.57-2.97; P=0.54) (**Figure 3E**).

Class-level longitudinal analyses were not performed, as no lipid class was associated with BP cross-sectionally.

### Sensitivity analyses

Sensitivity analyses did not materially alter the association between TG 58:9 FA 22:4 and systolic BP percentile (β in sensitivities ranging 7.05-8.21 vs 7.36 in the primary model) or longitudinal BP patterns (OR in sensitivities ranging 2.43-4.83 vs 2.78 in primary model for incident elevated BP and 1.90-2.34 vs 2.14 in primary model for persistent elevated BP vs stable-normal BP; and 2.57-4.60 vs 2.94 in primary model for incident elevated BP and 1.81-2.43 vs 2.27 in primary model for persistent elevated BP vs resolved elevated BP), with p-values<0.05 for all model specifications, except for persistent elevated BP vs resolved elevated BP when excluding children with overweight and obesity (**Figure S9**).

## Discussion

In this population-based study of preadolescent children, TG 58:9 FA 22:4 was positively associated with systolic BP percentile after lipidome-wide multiple-testing correction. Longitudinal analyses indicated that TG 58:9 FA 22:4 was higher in children with incident or persistent elevated BP than in those whose earlier elevated BP had resolved or those with stable-normal BP, supporting a closer relationship of this specific lipid with current BP elevation. Although comparable data are lacking in children, adult longitudinal findings show broadly concordant class-level directions. In the Strong Heart Family Study, higher baseline TG species were associated with incident hypertension; longitudinal increases in TG species were positively associated with systolic BP change [16].

TG 58:9 FA 22:4 is a long-chain (58 carbons in total), highly polyunsaturated (9 double bonds) TG in which MS/MS identified a 22-carbon FA moiety with four double bonds (FA 22:4), compatible with adrenic acid (22:4n-6) [32]. In our study, it was positively associated primarily with systolic BP, although a similar direction of association was observed across all BP outcomes at nominal P<0.05 and across all model specifications, including additional adjustment for fish and seafood consumption, which argues against habitual fish intake explaining this highly unsaturated TG signal. Among Chinese children from minority groups, hypertension was associated with higher long-chain, polyunsaturated TGs sharing either the total composition (TG 58:9) or the FA 22:4 residue, whereas a plasmalogen PE containing FA 22:4 was lower [21]. This suggests that associations of FA 22:4 with BP may depend on the lipid class and metabolic context in which it is incorporated, rather than reflecting a uniform effect. The magnitude of the association is potentially relevant at population level. A 1-SD higher TG 58:9 FA 22:4 corresponded to 7.36 systolic BP percentile, ∼ 0.31 SD of the systolic BP percentile distribution or ∼3 mmHg (based on the ratio of the SDs of systolic BP percentiles and mmHg in our population). In a pooled analysis of seven cohorts, each 1-unit higher childhood systolic BP z-score was associated with a 34% higher risk of fatal adult cardiovascular events [5]. The BP difference associated with this lipid is therefore of a magnitude that, if persistent, would be relevant for long-term cardiovascular risk.

Despite no evidence of enrichment for TG as a set, TG 58:9 FA 22:4 lay within a region of predominantly positive associations among longer-chain, more polyunsaturated TG species. Pediatric evidence was recently reported in a nested case-control lipidomic study of Chinese children, in which several long-chain, highly unsaturated TGs were elevated in children with higher BP [19]. Potential mechanisms linking TG-rich lipoprotein metabolism to BP include endothelial dysfunction and oxidative stress [33, 34], but this evidence relates to conventional hypertriglyceridemia and cannot be attributed to TG species.

Adiposity is an important potential determinant of both circulating TG profiles and BP. In children and adolescents, large, weakly unsaturated TG species were elevated in those suffering from overweight or obesity and decreased with greater reductions in BMI SDS during a lifestyle intervention [17]; while TG-related differences associated with hypertension were substantially attenuated after adjustment for BMI z score [19]. In contrast, our lead association (TG 58:9 FA 22:4 with systolic BP percentile) remained relatively stable after weight adjustment and after excluding children with overweight or obesity. This finding, together with the fact that TG 58:9 FA 22:4 is highly unsaturated, suggests that the observed association may reflect a pathway distinct from adiposity-driven TG accumulation; alternatively, adiposity may still contribute through a pathway that weight adjustment does not capture. Children with any elevated-BP pattern, including the resolved group, were more often overweight or obese than stable-normal children (26 to 31% vs 11%; **Table S1**), so adiposity history could contribute to the resolved-group signals.

Phospholipid species with 38 total carbons, mostly PC and PE carrying 20- and 22-carbon chains, were inversely associated with diastolic BP, consistent with the borderline PC pattern. As this set combines several lipid classes and was the only significant set, it should be considered hypothesis-generating.

This study has several strengths, including BP phenotyping via 5 repeated BP standardized measurements; an untargeted, lipid-wide approach across more than 1,200 species in a population-based, not clinically selected or obesity-restricted, pediatric cohort; rigorous correction for multiple testing via a permutation-based empirical family-wise error rate, including lipid-set inference by sample permutation, which accounts for the strong correlation among lipid species that is ignored by the commonly used preranked procedure [35]; extensive sensitivity analyses addressing confounding by adiposity, pubertal timing, seasonality, and comorbidity; and longitudinal BP phenotyping spanning early and later childhood.

This study also has limitations. The estimate for the lead lipid (β=7.36) is below the MDE (8.68), so smaller true effects may have gone undetected and the estimate may be inflated, as is common in discovery analyses. This is a single-cohort study without external validation. BP patterns rest on one oscillometric visit at each age, so some children may be misclassified and regression to the mean may contribute to the resolved group. Lipids were measured once, in preadolescence, and circulating lipid profiles are dynamic [36], so we cannot tell when the differences arose. The cohort is predominantly of European ancestry, limiting generalizability to other populations. Blood sampling was not standardized with respect to fasting status, which may contribute to interindividual variation in lipid species; adjustment for time of blood collection may only partially account for this source of variability. We acknowledge that our sex-interaction findings are exploratory and require replication, although plausibility is supported by lipidomic sexual dimorphism in preadolescents [37] and previous evidence that metabolic correlates of BP differ between males and females during adolescence [22].

From a translational perspective, our findings suggest that normalization of BP may coincide with normalization of BP-lipid-associated signatures. Previous life-course studies indicate that cardiovascular target-organ risk is substantially attenuated when elevated childhood BP resolves, although a small residual risk cannot be excluded [7, 38, 39]. However, longitudinal studies with repeated lipidomics and cardiovascular phenotyping are needed to determine whether persistent signatures exist among children whose earlier elevated BP has resolved.

In conclusion, our study found TG 58:9 FA 22:4 associated with systolic BP in preadolescents and was closely associated with current BP elevation. Replication in independent cohorts with repeated lipidomics is needed.

## Contributors

RA conceptualisation, accessed and verified the underlying data, formal analysis, visualization, writing–original draft, writing–review and editing. TSN supervision, funding acquisition, writing–review and editing. NG, ER and MV investigation, writing– review and editing. KB methods, writing–review and editing. MP supervision, funding acquisition, conceptualisation, accessed and verified the underlying data, writing– original draft, writing–review and editing. All authors contributed to the intellectual content and reviewed and approved the final manuscript for publication.

## Data sharing statement

The individual-level data underlying the findings of this study are not publicly available because they contain information from an ongoing birth cohort with active recruitment and follow-up, and because parts of the dataset are currently being used in other unpublished analyses and manuscripts. De-identified data relevant to the analyses presented in this manuscript may be made available from the corresponding author, Rossella Alfano, upon reasonable request after publication. Lipidome-wide results are available in Supplementary tables, and the analysis code used for the present study is publicly available on GitHub: https://github.com/Limburgsgeboortecohort/BP-lipidome. Requests should be directed to and will be considered for researchers with a methodologically sound proposal, subject to approval by the ENVIRONAGE study team and, where applicable, relevant ethical procedures.

## Data Availability

The individual-level data underlying the findings of this study are not publicly available because they contain information from an ongoing birth cohort with active recruitment and follow-up, and because parts of the dataset are currently being used in other unpublished analyses and manuscripts. De-identified data relevant to the analyses presented in this manuscript may be made available from the corresponding author, Rossella Alfano, upon reasonable request after publication. Requests should be directed to and will be considered for researchers with a methodologically sound proposal, subject to approval by the ENVIRONAGE study team and, where applicable, relevant ethical procedures.

## Acknowledgments

The researchers would like to thank ENVIR*ON*AGE participants, all the ENVIR*ON*AGE follow-up study researchers, midwives, and the staff of the maternity ward of East-Limburg Hospital in Genk.

This work was supported by a Methusalem grant, the Research Foundation Flanders (FWO, grant numbers 1516112 N, G.0873.11.N.10) and Kom op Tegen Kanker to the ENVIRONAGE birth cohort study. RA holds a BOF postdoctoral mandate (BOF26OWB16).

During the preparation of this work, the authors used ChatGPT and Claude in order to improve the language and readability of the manuscript. After using this tool, the authors reviewed and edited the content as needed and take full responsibility for the content of the publication.

## Supplemental Material

Supplemental Methods

Tables S1-S4

Figures S1-S9

