## Supplementary material for "Lipidome-wide analysis of concurrent and longitudinal blood pressure patterns in school-aged children"

Tables S2-S4 are available as separate excel files.

### Supplemental Methods

**Lipid extraction and LC–ESI–MS/MS analysis**

4 μL of plasma was mixed with 700 μL of water, 800 μl HCl(1M):CH3OH 1:8 (v/v), 900 μl CHCl3, 200 μg/ml of the antioxidant 2,6-di-tert-butyl-4-methylphenol (BHT; Sigma Aldrich), 3 μl of UltimateSPLASH™ ONE internal standard mix (#330820, Avanti Polar Lipids) and 3 μl of SphingoSPLASH™ I internal standard mix (#330734, Avanti Polar Lipids). After vortexing and centrifugation, the lower organic fraction was collected and evaporated using a Savant Speedvac spd111v (Thermo Fisher Scientific) at room temperature and the remaining lipid pellet was stored at - 20°C under argon. Just before mass spectrometry analysis, lipid pellets were reconstituted in 100% ethanol. Lipid species were analyzed by liquid chromatography electrospray ionization tandem mass spectrometry (LC-ESI/MS/MS) on a Nexera X2 UHPLC system (Shimadzu) coupled with hybrid triple quadrupole/linear ion trap mass spectrometer (6500+ QTRAP system; AB SCIEX). Chromatographic separation was performed on a XBridge amide column (150 mm × 4.6 mm, 3.5 μm; Waters) maintained at 35°C using mobile phase A [1 mM ammonium acetate in water-acetonitrile 5:95 (v/v)] and mobile phase B [1 mM ammonium acetate in water-acetonitrile 50:50 (v/v)] in the following gradient: 0–6 min, 0–6% B; 6–10 min, 6–25% B; 10–11 min, 25–98% B; 11–13 min, 98–100% B; 13–19 min, 100% B; and 19–24 min, 0% B. The flow rate was 0.7 mL/min until 13 min and was subsequently increased to 1.5 mL/min. Sphingomyelins (SMs), cholesteryl esters (CEs), ceramides (Cers), dihydroceramides, hexosylceramides (HexCers), and lactosylceramides (LacCers; annotated as Hex2Cers in the analytical output), were measured in positive-ion mode using product ions with mass-to-charge ratios of 184.1, 369.4, 264.4, 266.4, 264.4, and 264.4, respectively. TGs, DGs, and monoacylglycerols were measured in positive-ion mode using neutral losses corresponding to individual fatty acyl moieties. PCs, lysophosphatidylcholines (LPCs), PEs, lysophosphatidylethanolamines (LPEs), phosphatidylglycerols (PGs), phosphatidylinositols (PIs), and phosphatidylserines were measured in negative-ion mode using fatty acyl fragment ions. Lipid quantification was performed by scheduled multiple reactions monitoring (MRM), the transitions being based on the neutral losses or the typical product ions as described above. The instrument parameters were as follows: Curtain Gas = 35 psi; Collision Gas = 8 a.u. (medium); IonSpray Voltage = 5500 V and −4,500 V; Temperature = 550°C; Ion Source Gas 1 = 50 psi; Ion Source Gas 2 = 60 psi; Declustering Potential = 60 V and −80 V; Entrance Potential = 10 V and −10 V; Collision Cell Exit Potential = 15 V and −15 V. The following fatty acyl (FA) moieties were taken into account for the lipidomic analysis: 14:0, 14:1, 16:0, 16:1, 16:2, 18:0, 18:1, 18:2, 18:3, 20:0, 20:1, 20:2, 20:3, 20:4, 20:5, 22:0, 22:1, 22:2, 22:4, 22:5 and 22:6 except for TGs, which considered: 16:0, 16:1, 18:0, 18:1, 18:2, 18:3, 20:3, 20:4, 20:5, 22:2, 22:3, 22:4, 22:5, 22:6. Peak integration was performed with the MultiQuantTM software version 3.0.3. Lipid species signals were corrected for isotopic contributions (calculated with Python Molmass 2019.1.1) and were quantified based on internal standard signals and adheres to the guidelines of the Lipidomics Standards Initiative (LSI) (level 2 type quantification as defined by the LSI).

**Covariates**

***Main covariates***

At the follow-up visit at 9-12 years, trained fieldworkers measured body anthropometrics of the children including height (to the nearest centimeter) and weight (to the nearest 0.1 kilogram), from which BMI was derived as kg/m^2^. Detailed questionnaires are filled out by the mothers to obtain household and lifestyle information. Household smoke status during the follow-up period was collected (absent, when none of the parents smoked in house, or present when at least one parent smoked in house). The highest educational attainment of either parent was categorized as low when the highest completed level was secondary education or lower, and as high when at least one parent had completed tertiary education, including bachelor, graduate, higher vocational education, master, or doctoral education. Based on the date of follow-up, a seasonal scale was calculated (winter from December to February, spring from March to May, summer from June to August, autumn from September to November). Average daily physical activity and screen time were derived from parental reports of the time the child typically spent performing these activities. Screen time included time spent watching television or using computers, tablets, smartphones, or other screen-based devices. Physical activity comprised the reported time spent in active play, sports, or other physical activities. Information was collected separately for weekdays and weekend days, and a weekly weighted average was calculated.

The time of blood collection was recorded as the hour of the day. Plasma storage duration was calculated as the interval between blood collection and lipidomic analysis. Ethnicity (non-European if the country of origin of 3 or more grandparents was non-European, otherwise European) was collected via a questionnaire self-completed by mothers at the child's birth.

***Additional covariates considered in sensitivity analyses***

Children with overweight and obesity who reported cardiometabolic conditions and corticosteroid use were excluded in separate sensitivity analyses to assess whether these conditions or treatments disproportionately influenced the observed associations. Overweight and obesity were classified according to age- and sex-specific International Obesity Task Force criteria. Cardiometabolic conditions and corticosteroid use were reported via questionnaires filled out by mothers.

Average daily ambient temperature at the child’s residential address on the day of examination was obtained from IRCEL-CELINE. Temperature was available for examinations conducted up to 2023 but could not be retrieved for a subset of examinations conducted in 2024, and was therefore evaluated in sensitivity analyses among participants with available data.

Pubertal development was assessed using a parent-reported adaptation of the Pubertal Development Scale (PDS). The scale included growth spurt, body-hair growth and skin changes in all children; voice change and facial-hair growth in boys; and breast development and menarche in girls. Items were scored from 1, indicating that development had not started, to 4, indicating completed development; “I do not know” responses were treated as missing. Menarche was scored as 1 for no and 4 for yes. Sex-specific scores were calculated as the mean of the five applicable items when at least three items were available. Pubertal development was not included in the primary adjustment set because the cohort was predominantly pre-/early-pubertal, with relatively limited variation in PDS scores; models additionally adjusted for PDS were therefore examined as a sensitivity analysis.

Children’s fish and seafood consumption during the previous month was assessed via a self-completed food frequency questionnaire. Frequency of consumption was assessed separately for non-fat fish, half-fat fish, fatty fish, and shellfish. Frequency responses ranged from never to more than once daily and were combined into a total fish and seafood consumption variable.

**Multiple-testing correction and empirical family-wise error estimation**

To account for multiple testing while preserving dependence among lipid species, empirical family-wise error (FWER)-adjusted P values ($P_{\mathrm{FWER}}$) were calculated using a single-step maximum-statistic permutation procedure. Nuisance covariates - i.e., all covariates included in the model other than the lipid species under evaluation - were handled using the Freedman-Lane residual-permutation approach. Residuals from the corresponding covariate-only model were permuted and added to its fitted values. For each of 50,000 permutations, associations with all 1,232 lipid species were evaluated and the maximum absolute t-statistic was retained. For each lipid species, $P_{\mathrm{FWER}}$ was calculated by comparing its observed absolute t-statistic with the permutation distribution of maximum absolute t-statistics. This procedure is based on the Westfall-Young framework; related permutation-based approaches have previously been applied in metabolomic and lipidomic epidemiology. Lipidome-wide significance was defined as P_FWER_<0.05

**Minimum detectable effect calculation**

We estimated the minimum detectable effect (MDE) using a simulation-based power analysis. Analyses were conducted separately for systolic and diastolic BP percentiles and for the sensitivity basic model (reduced covariates set, N=195) and in the primary model (N=186). For each outcome and model, a covariate-only linear model was fitted and its fitted values and residuals retained. Candidate effect sizes were evaluated over a grid of effect sizes (0–11 percentile points per 1-SD higher lipid), and 50,000 simulated datasets were generated for each effect size. Synthetic outcomes were constructed as the fitted covariate component plus the imposed lipid effect and an independently bootstrapped model residual. TG 58:9 FA 22:4, as the species with the most significant association in the primary analysis, was used as an index lipid to provide an empirically observed standardized lipid distribution. In each simulated dataset, the lipid-BP model was refitted and an association was considered detected when its nominal P value was below the outcome- and model-specific threshold corresponding to P_FWER_=0.05. These thresholds were derived from the 50,000-permutation Freedman-Lane maxT distribution of the corresponding lipidome-wide analysis and therefore accounted for dependence among the 1,232 lipid tests. The MDE was defined as the effect size corresponding to 80% power, estimated by linear interpolation between adjacent grid points. Standardized MDEs were obtained by dividing the MDE by the SD of the corresponding BP percentile outcome.

The MDEs were 8.51 and 7.30 percentile points for systolic and diastolic BP, respectively, in the basic model, and 8.68 and 7.46 percentile points in the primary model, indicating the power estimate was not highly sensitive to the choice of covariate set (**Figure S3**). Because the index lipid was resampled independently of the covariates, these values should be interpreted as approximate MDEs for lipid species with distributions similar to the index lipid.

**Departure from linearity and model assumptions of the lipid species significant in the lipidome-wide association study**

For lipid species meeting the empirical FWER threshold in the main lipidome-wide association models, departures from linearity and model assumptions were assessed as follows.

Departure from linearity was explored using restricted cubic splines with three knots placed at the 10th, 50th and 90th percentiles of the lipid distribution, with the median concentration used as the reference and nonlinearity assessed by testing the nonlinear spline component.

Model assumptions were assessed using Q-Q plots of residuals, residual-versus-fitted plots, and Cook’s distance. Robustness to heteroscedasticity was assessed using HC3 heteroscedasticity-consistent standard errors.

### ****Table S1****

**Characteristics by BP patterns of 177 ENVIR*ON*AGE participants with available BP measurements at 4-6 years and at 9-12 years**

| **Characteristic** | **Longitudinal BP patterns (BP category transitions between 4-6 years and 9-12 years)** | | | | **P value** |
| --- | --- | --- | --- | --- | --- |
|  | **Stable normal (n=100)** | **Resolved elevated BP (n=28)** | **Incident elevated BP (n=23)** | **Persistent elevated BP (n=26)** |  |
| **9-12 years** | | | | | |
| Child sex, boys | 45 (45.0) | 11 (39.3) | 13 (56.5) | 10 (38.5) | 0.563 |
| Age, years | 10.11 (0.81) | 10.03 (0.86) | 9.68 (0.68) | 10.29 (0.84) | 0.060 |
| Ethnicity, non-European | 6 (6.0) | 3 (10.7) | 2 (8.7) | 1 (3.8) | 0.736 |
| BMI, kg/m² | 17.26 (2.58) | 18.11 (2.70) | 17.93 (2.77) | 18.71 (2.85) | 0.066 |
| Weight, Kg | 34.68 (7.25) | 36.14 (7.25) | 35.49 (7.42) | 38.28 (8.71) | 0.178 |
| Height, cm | 141.29 (7.33) | 140.87 (7.52) | 140.24 (7.71) | 142.29 (7.52) | 0.799 |
| Overweight or obese | 11 (11.0) | 8 (28.6) | 6 (26.1) | 8 (30.8) | **0.029** |
| Highest parental educational attainment, low | 19 (19.2) | 4 (14.8) | 8 (34.8) | 4 (15.4) | 0.263 |
| Household smoking, yes | 23 (23.5) | 4 (15.4) | 6 (26.1) | 6 (23.1) | 0.806 |
| Average daily screen time, h/day | 3.67 (2.32) | 3.35 (1.68) | 3.41 (2.01) | 4.12 (2.53) | 0.599 |
| Average daily physical activity, h/day | 3.82 (2.38) | 4.41 (2.58) | 4.02 (2.26) | 5.03 (2.42) | 0.132 |
| Pubertal Development Scale score | 1.33 (0.48) | 1.37 (0.36) | 1.25 (0.38) | 1.44 (0.48) | 0.482 |
| Season of follow-up, n (%) |  |  |  |  | 0.951 |
| *Winter* | 25 (25.0) | 6 (21.4) | 4 (17.4) | 5 (19.2) |  |
| *Spring* | 22 (22.0) | 8 (28.6) | 4 (17.4) | 6 (23.1) |  |
| *Summer* | 34 (34.0) | 7 (25.0) | 8 (34.8) | 9 (34.6) |  |
| *Autumn* | 19 (19.0) | 7 (25.0) | 7 (30.4) | 6 (23.1) |  |
| Average temperature, °C | 12.35 (6.09) | 11.52 (5.96) | 12.75 (5.86) | 10.45 (4.88) | 0.485 |
| Fish and seafood consumption, time per week | 2.02 (2.07) | 1.77 (1.01) | 1.75 (0.95) | 1.62 (0.97) | 0.713 |
| Collection time, hour | 11.93 (2.20) | 12.89 (2.14) | 12.89 (1.98) | 12.21 (2.03) | 0.084 |
| Plasma storage duration, years | 3.10 (0.95) | 2.97 (1.00) | 3.19 (0.75) | 3.31 (0.93) | 0.593 |
| Systolic BP, mmHg | 102.71 (7.08) | 103.46 (5.30) | 116.26 (5.88) | 119.23 (8.34) | **<0.001** |
| Diastolic BP, mmHg | 62.94 (5.41) | 63.19 (5.39) | 70.99 (5.45) | 68.54 (7.86) | **<0.001** |
| Mean arterial pressure, mmHg | 78.85 (5.27) | 79.30 (4.46) | 89.10 (3.82) | 88.82 (6.29) | **<0.001** |
| Systolic BP percentile | 58.47 (22.60) | 61.43 (17.81) | 92.52 (7.08) | 94.46 (4.79) | **<0.001** |
| Diastolic BP percentile | 56.39 (18.86) | 58.54 (19.44) | 80.83 (14.59) | 71.50 (21.38) | **<0.001** |
| BP category |  |  |  |  | **<0.001** |
| Normal BP | 100 (100.0) | 28 (100.0) | 0 (0.0) | 0 (0.0) |  |
| Elevated BP | 0 (0.0) | 0 (0.0) | 8 (34.8) | 10 (38.5) |  |
| Stage 1 hypertension | 0 (0.0) | 0 (0.0) | 11 (47.8) | 10 (38.5) |  |
| Stage 2 hypertension | 0 (0.0) | 0 (0.0) | 4 (17.4) | 6 (23.1) |  |
| **4-6 years** | | | | | |
| Systolic BP percentile, percentile points | 61.07 (21.62) | 87.82 (12.30) | 64.13 (19.79) | 94.23 (4.33) | **<0.001** |
| Diastolic BP percentile, percentile points | 53.76 (20.78) | 72.54 (25.90) | 59.35 (19.31) | 71.19 (28.18) | **<0.001** |

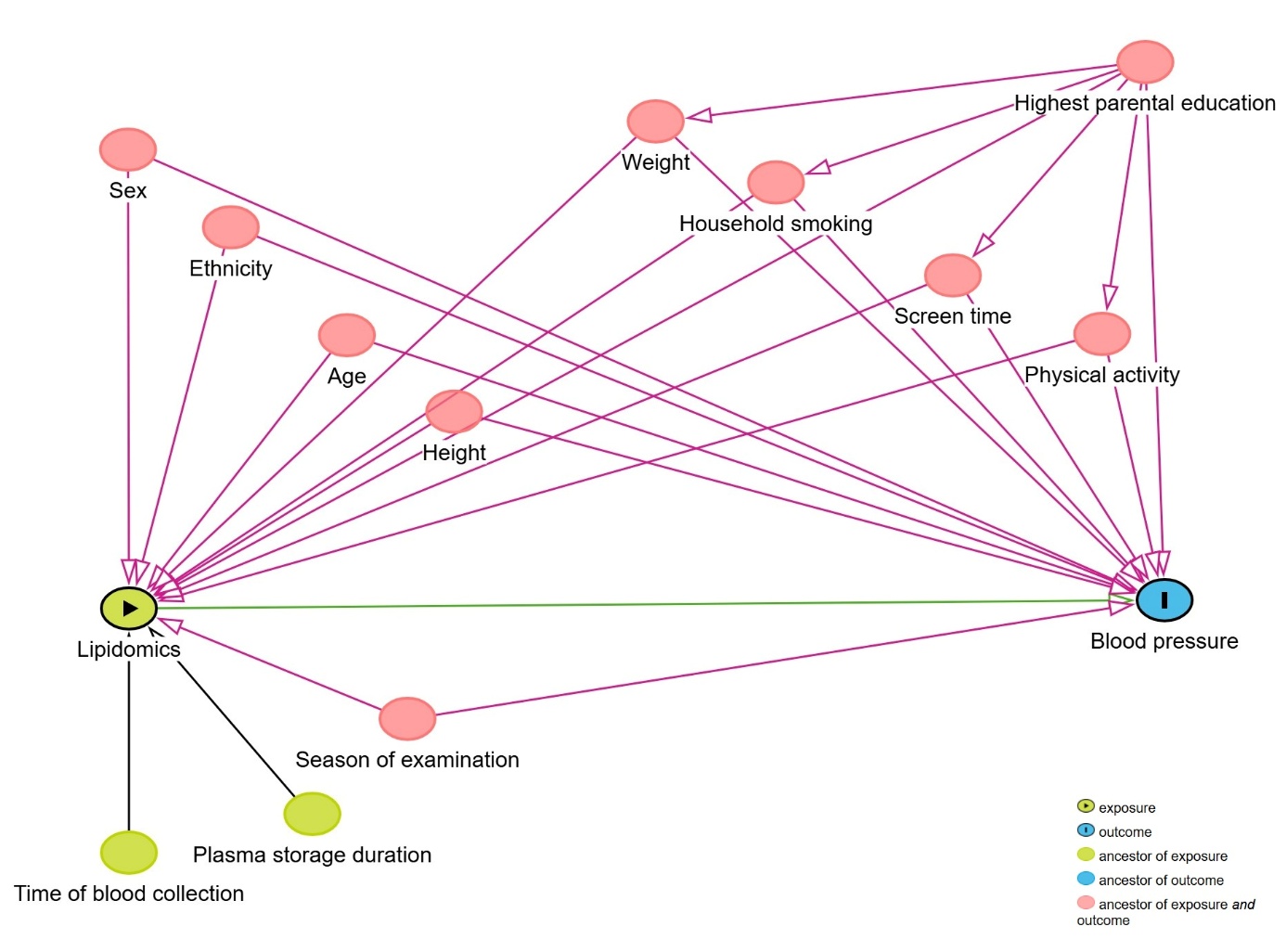

### ****Figure S1****

**Directed acyclic graph (DAG) informing covariate selection for the association between plasma lipidomics and blood pressure (BP) at 9-12 years. The DAG represents hypothesized relationships among plasma lipid concentrations, blood pressure (BP), and potential common determinants of both. Child age, sex, ethnicity, height, weight, highest parental educational attainment, household smoking, physical activity, screen time, and season of examination were considered potential confounders of the lipidomics-BP association. Time of blood collection and plasma storage duration were identified as temporal and preanalytical covariates.**

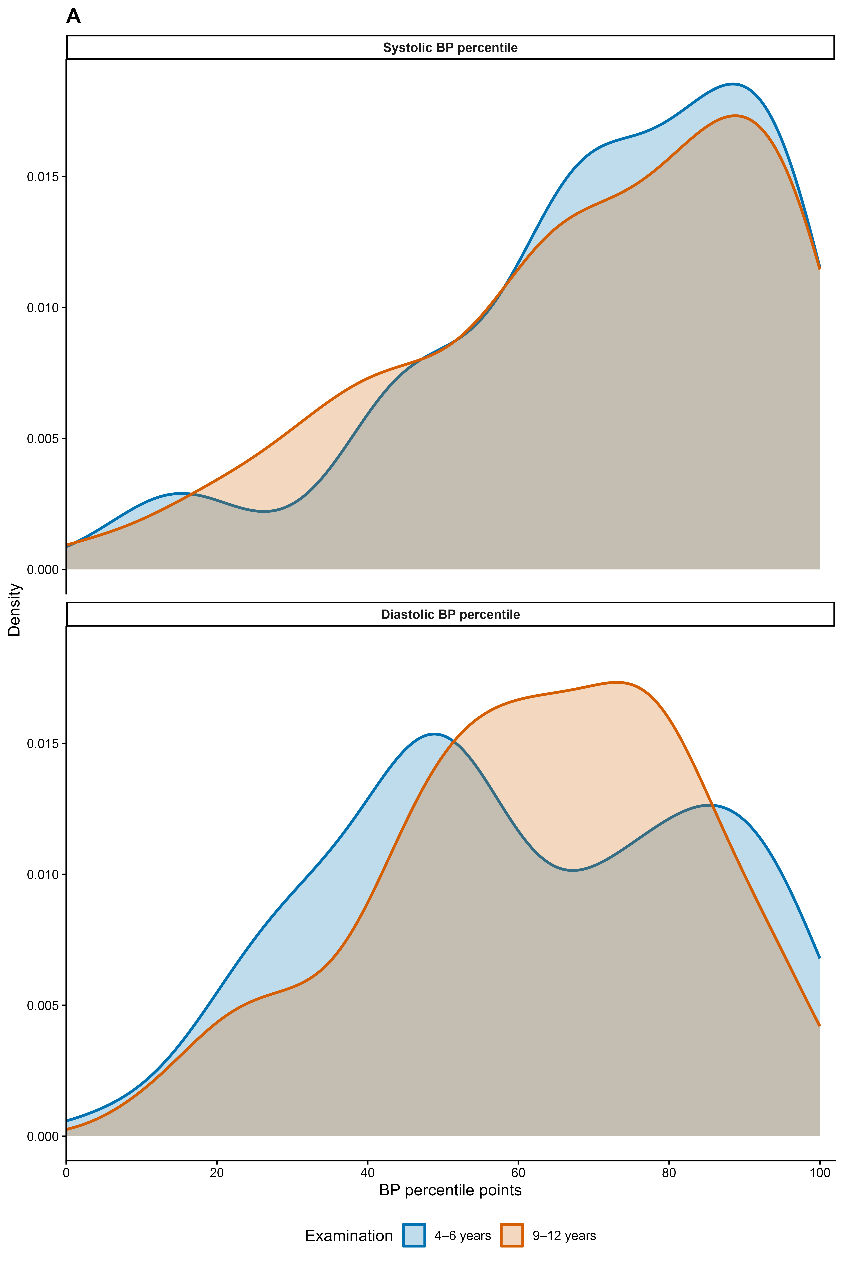

### Figure S2

**Blood pressure percentile distributions across childhood.** Distributions of systolic and diastolic BP percentiles at the childhood examinations conducted at 4-6 and 9-12 years of age
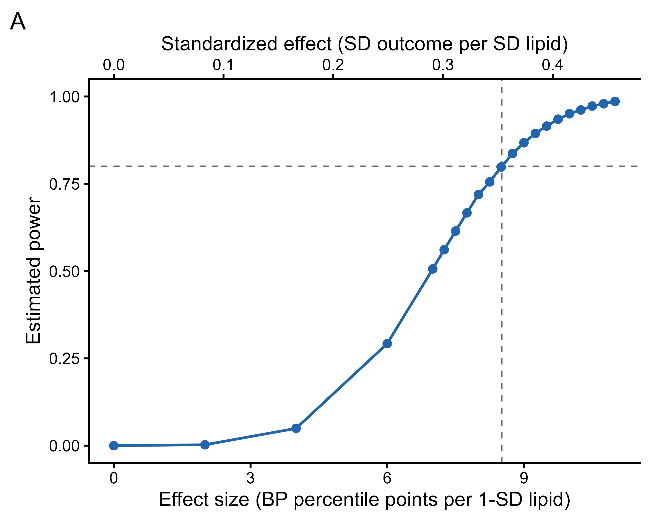

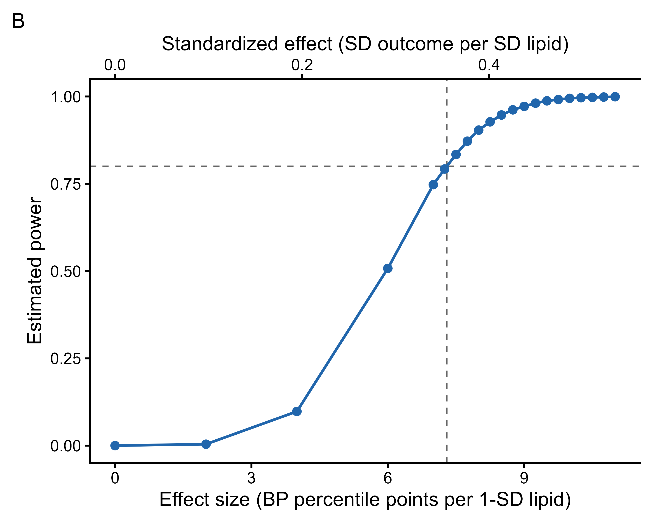

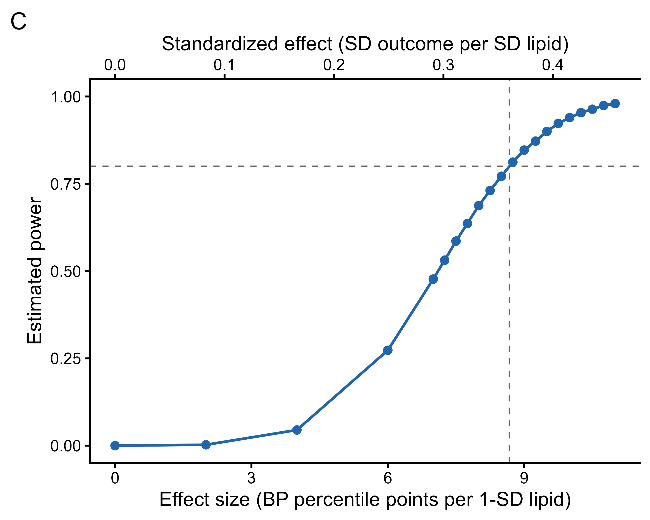

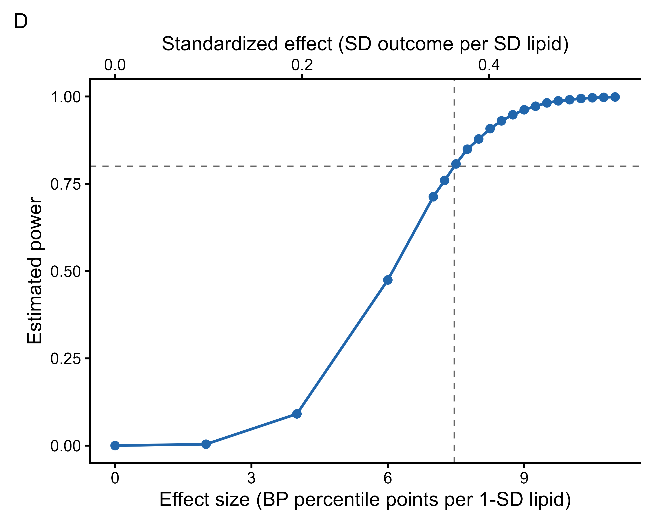

### Figure S3

**Simulated power to detect a lipid-BP association at the study-wide significance threshold.** Power curves are shown for systolic (A) and diastolic (B) BP percentile in the sensitivity basic model (N = 195), adjusted for child age, sex, height, ethnicity, season of examination, time of blood collection and sample storage time, and for systolic (C) and diastolic (D) BP percentile in the primary model (N = 186), additionally adjusted for child weight, average daily physical activity, average daily screen time, highest parental educational attainment and household smoking. The y-axis shows the estimated power. The lower x-axis shows the effect size as BP percentile points per 1-standard-deviation (SD) higher log₂-transformed lipid concentration, and the upper x-axis the corresponding standardized effect (SD of BP percentile per SD of lipid). In all panels, the horizontal dashed line indicates 80% power and the vertical dashed line the corresponding minimum detectable effect (MDE). Details of the MDE estimation can be found in the **Supplemental Methods**.

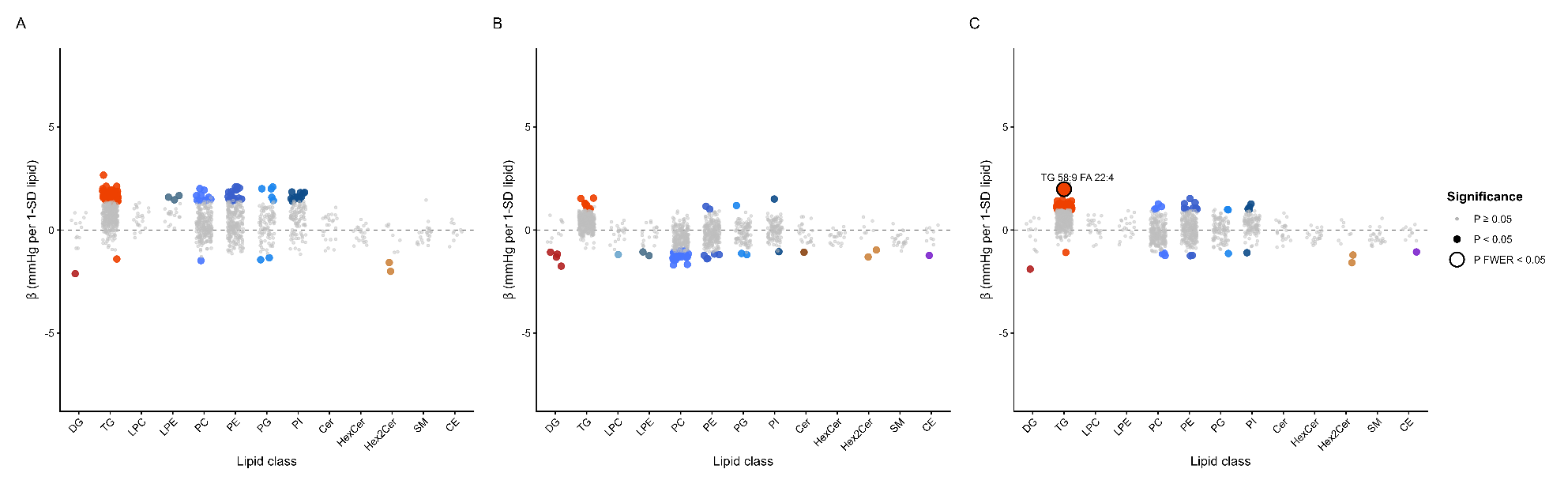

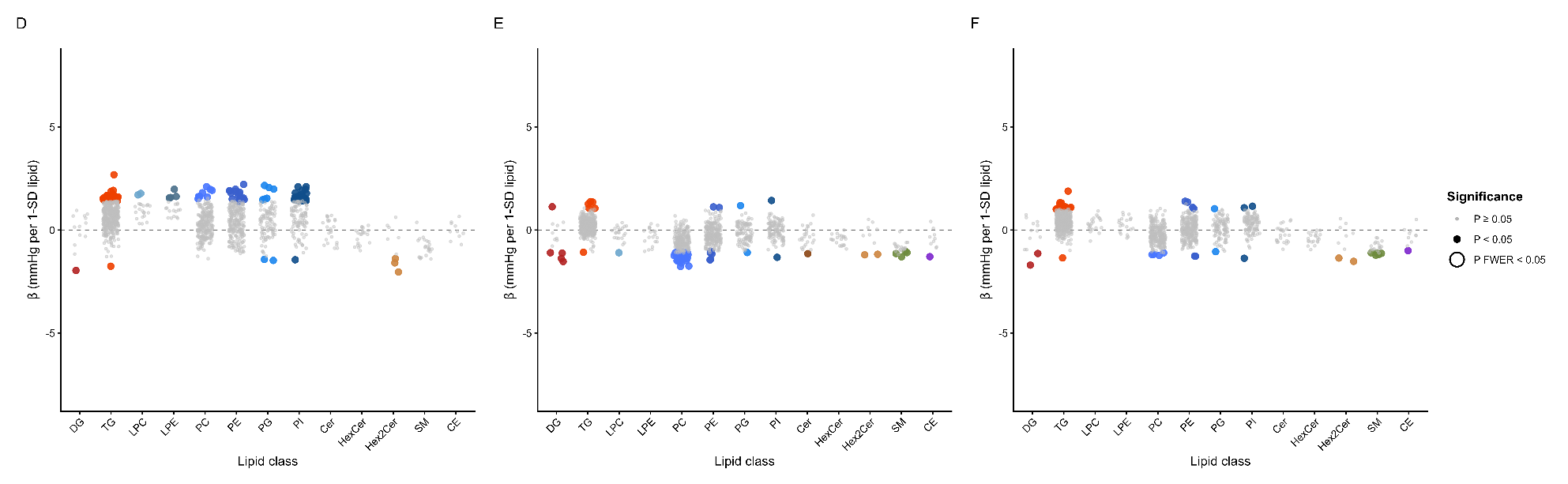

### Figure S4

**Associations of individual plasma lipid species with blood pressure expressed as mmHg.** Regression coefficients for associations of individual lipid species in sensitivity basic model (**A-C**) and the primary model (**D-F**) with (**A,D**) systolic and (**B,E**) diastolic and (**C,F**) mean arterial blood pressure (BP) expressed as mmHg are presented according to lipid class. Coefficients represent the difference in blood pressure, in mmHg, associated with a 1-standard-deviation higher log₂-transformed lipid concentration. Grey points represent P≥0.05, small coloured points represent nominal P<0.05, and enlarged coloured points with a black outline represent $P_{\mathrm{FWER}}$<0.05. Basic models included 195 children and were adjusted for child age, sex, height, ethnicity, season of examination, time of blood collection and plasma storage duration. Primary models included 186 children and were adjusted for the same covariates of the basic model in addition to child weight, average daily physical activity, average daily screen time, highest parental educational attainment and household smoking. The horizontal dashed line indicates the null value.

CE= cholesteryl ester; Cer= ceramide; DG= diacylglycerol; HexCer= hexosylceramide; Hex2Cer= dihexosylceramide; LPC= lysophosphatidylcholine; LPE= lysophosphatidylethanolamine; PC= phosphatidylcholine; PE= phosphatidylethanolamine; PG= phosphatidylglycerol; PI= phosphatidylinositol; SM= sphingomyelin; TG= triacylglycerol

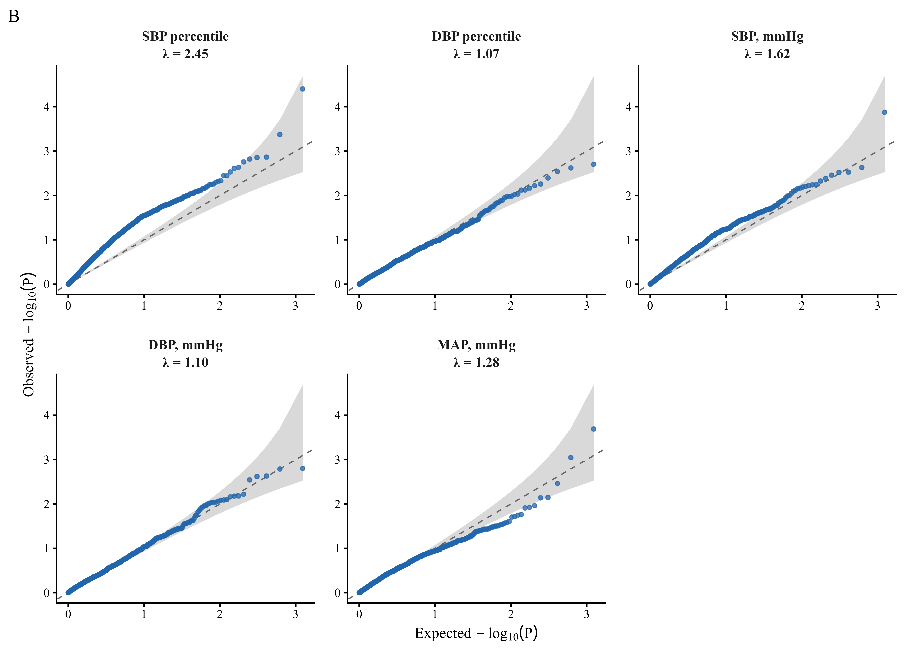

### Figure S5

**Quantile-quantile plots for the lipid-wide association analyses.** Observed P-value distributions are plotted against their expected distributions for systolic and diastolic BP percentiles and for systolic BP (SBP), diastolic BP (DBP) and mean arterial pressure (MAP) expressed in mmHg. Primary models included 186 children and were adjusted for child age, sex, height, ethnicity, weight, average daily physical activity, average daily screen time, highest parental educational attainment and household smoking, season of examination, time of blood collection and plasma storage duration. Shaded areas represent pointwise 95% confidence envelopes under independently distributed P values. The diagonal dashed line indicates the null distribution.

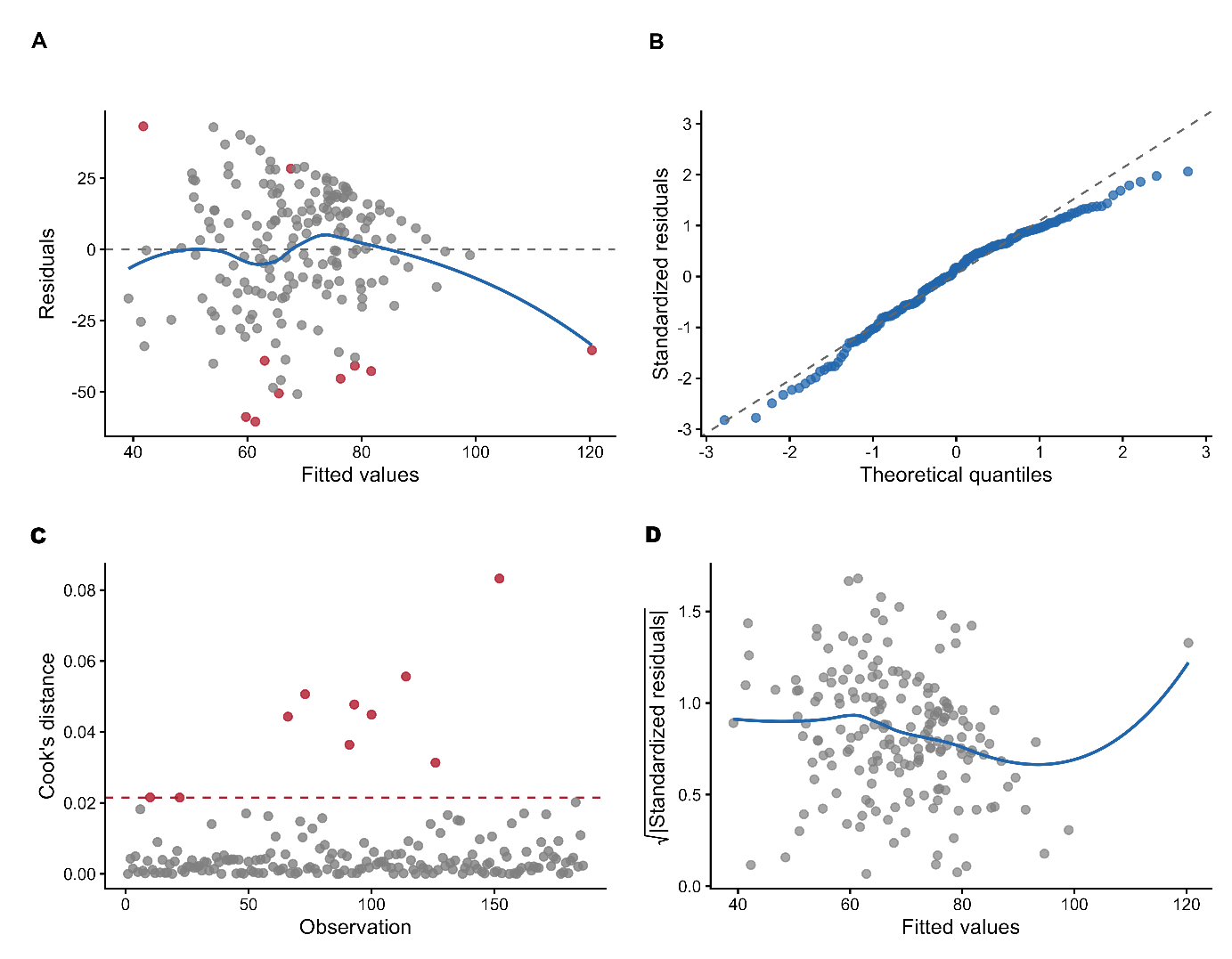

### Figure S6

**Diagnostic assessment of the association between TG 58:9 FA 22:4 and systolic blood pressure (BP) percentile.** Diagnostics are presented for the primary model (adjusted for child age, sex, ethnicity, height, weight, season of examination, time of blood collection, plasma storage duration, highest parental educational attainment, household smoking, average daily physical activity and average daily screen time), including 186 children**. (A)** Residuals plotted against fitted values. The horizontal dashed line indicates zero, and the blue line represents a locally estimated scatterplot smoothing curve. Red points identify observations with Cook’s distance greater than 4/n. **(B)** Quantile-quantile plot of standardized residuals; the dashed line represents the expected distribution under normality. **(C)** Cook’s distance for each observation. The horizontal red dashed line indicates the prespecified 4/n threshold, and red points indicate observations exceeding this threshold. **(D)** Scale-location plot of the square root of the absolute standardized residuals against fitted values; the blue line represents a locally estimated scatterplot smoothing curve. The association was additionally evaluated using HC3 heteroscedasticity-consistent standard errors and after excluding observations exceeding the Cook’s-distance threshold.

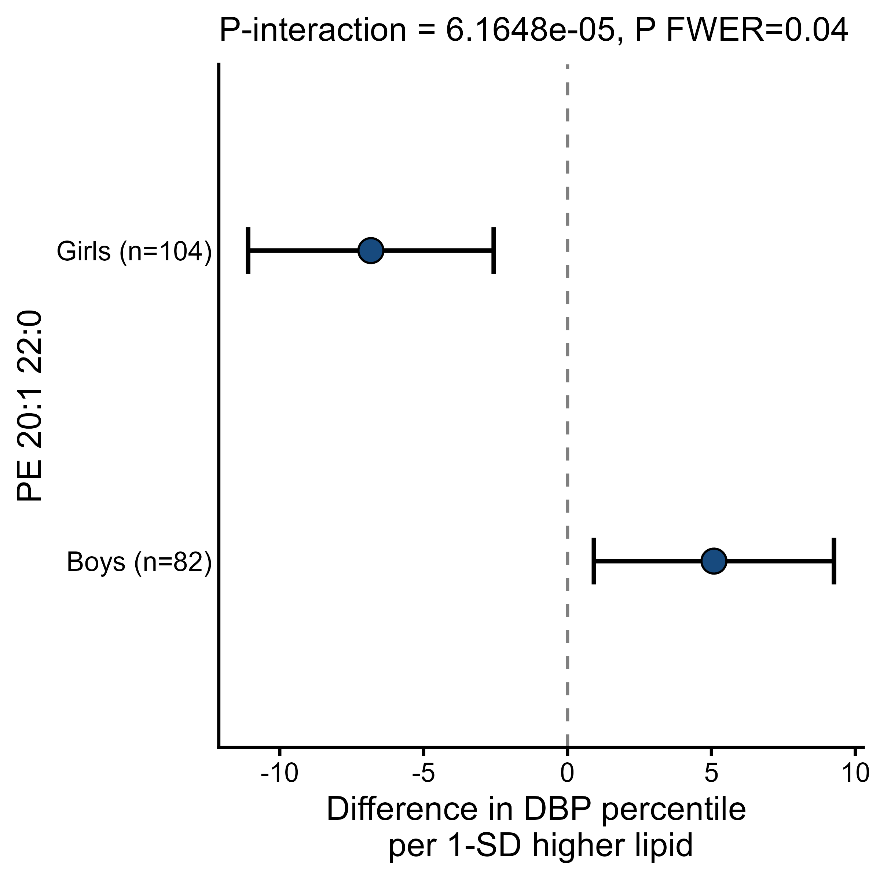

### Figure S7

**Sex-specific association of PE 20:1 22:0 with diastolic blood pressure percentile. Sex-stratified associations of standardized log₂-transformed PE 20:1 22:0 concentration with diastolic blood pressure (DBP) percentile in 186 children. Points represent the estimated difference in DBP percentile associated with a 1-SD higher lipid concentration, and horizontal bars represent 95% confidence intervals. Estimates were derived from the primary model including a lipid-by-sex interaction term and adjusted for child age, height, weight, ethnicity, highest parental educational attainment, household smoking, average daily physical activity, average daily screen time, season of examination, time of blood collection, and plasma storage duration. The dashed vertical line indicates the null value. The nominal interaction P value was 6.52×10⁻⁵ and the lipidome-wide family-wise error rate (FWER)–adjusted interaction P value was 0.044.**

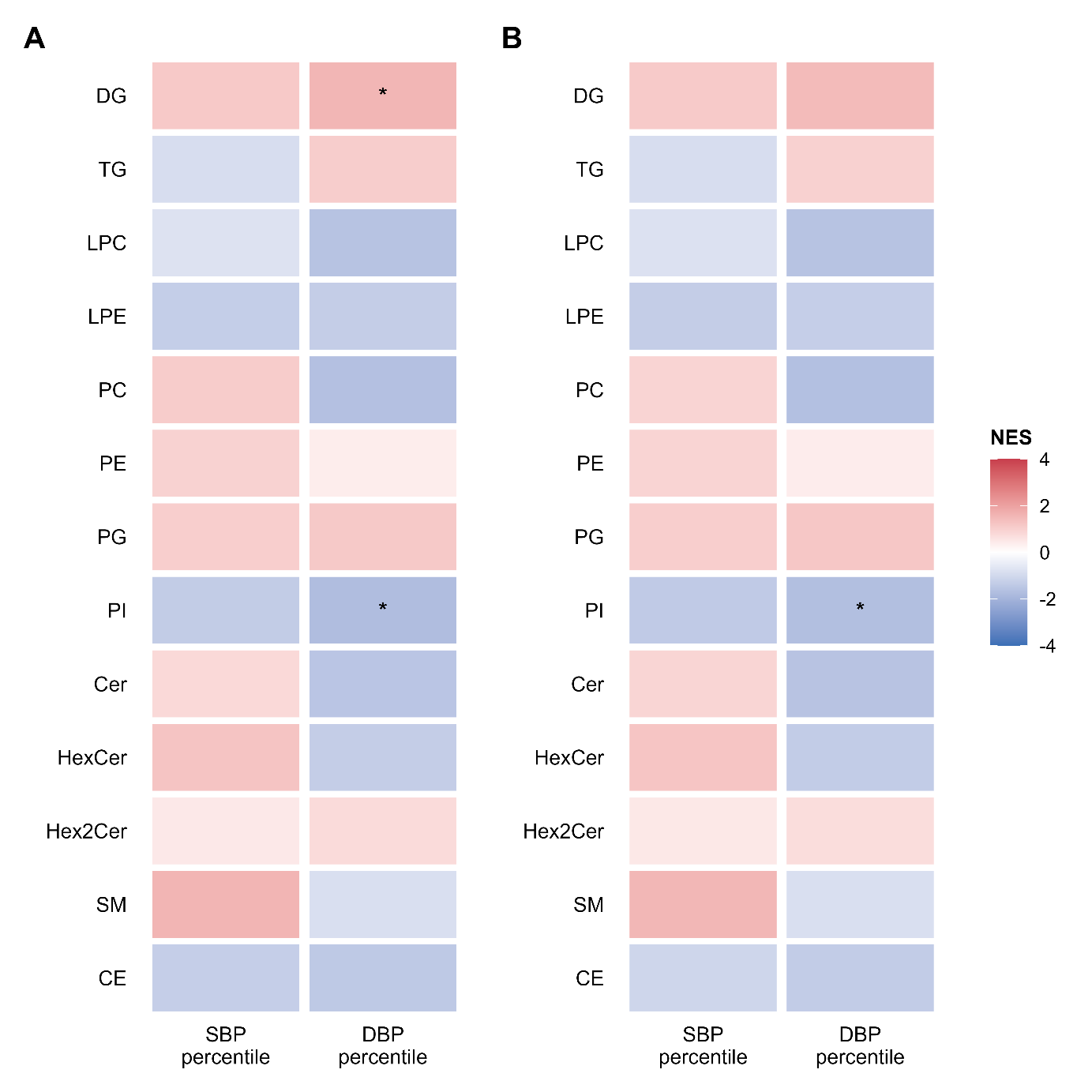

### Figure S8

**Lipid-class enrichment of sex-specific associations between plasma lipids and blood pressure.**

**Lipid-set enrichment analysis (LSEA) was applied to the signed lipid × sex interaction statistics for systolic (SBP) and diastolic (DBP) blood pressure percentiles. (A) Primary model. (B) Primary model additionally adjusted for pubertal development score (PDS). The primary model was adjusted for child age, sex, height, and ethnicity, season of examination, time of blood collection, and plasma storage duration, child weight, average daily physical activity, average daily screen time, highest parental educational attainment, and household smoking. Cell colour represents the normalized enrichment score (NES), with positive and negative values indicating enrichment toward positive and negative lipid × sex interaction coefficients, respectively. Asterisks indicate FDR-adjusted significance: *P<0.05; **FDR P<0.05. DG=diacylglycerol; TG=triacylglycerol; LPC=lysophosphatidylcholine; LPE=lysophosphatidylethanolamine; PC=phosphatidylcholine; PE=phosphatidylethanolamine; PG=phosphatidylglycerol; PI=phosphatidylinositol; CE=cholesteryl ester.**

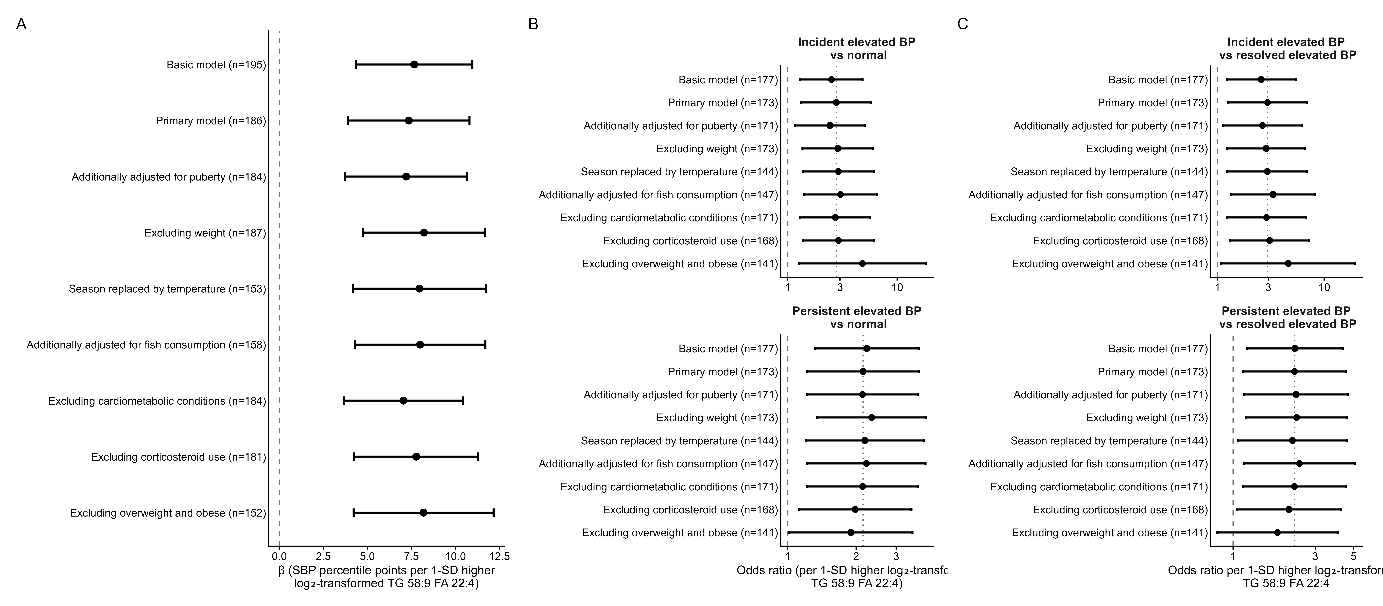

### Figure S9

**Sensitivity analyses of the association between TG 58:9 FA 22:4 and systolic blood pressure (BP) percentile and longitudinal BP patterns.** (**A**) Points represent adjusted differences in systolic BP percentile associated with a 1-SD higher log₂-transformed TG 58:9 FA 22:4 concentration; horizontal lines represent 95% confidence intervals. (**B**) Points represent odds ratios for incident and persistent elevated BP (versus stable-normal BP) and (**C**) for incident and persistent elevated BP (vs resolved elevated BP) associated with a 1-SD higher log₂-transformed TG 58:9 FA 22:4 concentration, from multinomial logistic regression; horizontal lines represent 95% confidence intervals, shown on a logarithmic x-axis. The basic model was adjusted for child age, sex, height, and ethnicity, season of examination, time of blood collection, and plasma storage duration; the primary model was additionally adjusted for child weight, average daily physical activity, average daily screen time, highest parental educational attainment, and household smoking. Sensitivity analyses repeated the primary model after: additionally adjusting for the Pubertal Development Scale score; excluding child weight; replacing season of examination with average daily temperature on the day of examination; additionally adjusting for fish consumption; excluding children with reported cardiometabolic conditions; excluding children with reported corticosteroid use; and excluding children with overweight or obesity. The vertical dashed line indicates the null value; the vertical dotted line indicates the point estimate of the primary model. Sample sizes for each analysis are shown in parentheses.
